# Genome-wide pleiotropy analysis of coronary artery disease and pneumonia identifies shared immune pathways

**DOI:** 10.1101/2021.07.05.21260028

**Authors:** Zhi Yu, Seyedeh M. Zekavat, Sara Haidermota, Rachel Bernardo, Peter Libby, Hilary Finucane, Pradeep Natarajan

**Affiliations:** Program in Medical and Population Genetics, Broad Institute of Harvard and MIT, Cambridge, MA; Cardiovascular Research Center, Massachusetts General Hospital, Boston, MA; Yale School of Medicine, New Haven, CT; Department of Medicine, Harvard Medical School, Boston, MA; Division of Cardiovascular Medicine, Brigham and Women’s Hospital, Boston, MA; Analytic and Translational Genetics Unit, Massachusetts General Hospital, Boston, MA

**Author notes:** Please address correspondence to: Pradeep Natarajan, MD MMSc, 185 Cambridge Street, CPZN 3.184, Boston, MA 02114, Twitter: @pnatarajanmd.

## Abstract

Coronary artery disease (CAD) remains the leading cause of death despite scientific advances. Elucidating shared CAD/pneumonia pathways may reveal novel insights regarding CAD pathways. We performed genome-wide pleiotropy analyses of CAD and pneumonia, examined the causal effects of the expression of genes near independently replicated SNPs and interacting genes with CAD and pneumonia, and tested interactions between disruptive coding mutations of each pleiotropic gene and smoking status on CAD and pneumonia risks. Identified pleiotropic SNPs were annotated to *ADAMTS7* and *IL6R*. Increased *ADAMTS7* expression across tissues consistently showed decreased risk for CAD and increased risk for pneumonia; increased *IL6R* expression showed increased risk for CAD and decreased risk for pneumonia. We similarly observed opposing CAD/pneumonia effects for *NLRP3*. Reduced *ADAMTS7* expression conferred a reduced CAD risk without increased pneumonia risk only among never-smokers. Genetic immune-inflammatory axes of CAD linked to respiratory infections implicate *ADAMTS7* and *IL6R*, and related genes.

## Introduction

Despite continued advances, coronary artery disease (CAD) remains the leading cause of mortality and disability-adjusted life-years (DALYs) worldwide^1^. Acute respiratory infections such as pneumonia, from influenza, SARS-CoV-2, and other pathogens, are common among individuals with chronic CAD^2, 3^ and can precipitate CAD events or exacerbate CAD symptoms^4^, and influenza vaccination reduces CAD risk^5, 6 7^. Such observations implicate the operation of immune-related CAD pathways. Elucidating shared pathways may reveal novel insights toward addressing the sizable persistent CAD risk in the population.

Prior human genetic studies identified interleukin 6 receptor (*IL6R*) as a putative causal factor for CAD^8, 9^, bolstered by the CANTOS (Canakinumab Anti-Inflammatory Thrombosis Outcomes Study) trial showing that inhibition of interleukin 1B (*IL1B*), a cytokine immediately upstream of IL-6, reduces CAD risk^10^. However, key questions persist: since *IL1B* inhibition was associated with increased infection risk, does this extend to other targets in the *IL1B* signaling pathway? Are there other inflammatory pathways implicating CAD and pneumonia? Can environmental factors influence the CAD and pneumonia risk associated with these pathways?

Here, we leveraged the pleiotropy between CAD and pneumonia to dissect jointly the unaddressed axis of CAD linked to pneumonia. We started with genome-wide pleiotropy analyses of CAD and pneumonia using summary statistics from CARDIoGRAMplusC4D and FinnGen R4. Then we examined the causal effects of genes near the pleiotropic loci that were independently replicated, as well as other closely interacting genes according to the STRING database of protein-protein interactions, on CAD and pneumonia through Mendelian randomization (MR). Lastly, we examined for interactions between those genes and smoking status on incident CAD and pneumonia risks in the UK Biobank. Our observations support genetic immune-inflammatory axes of CAD linked to respiratory infections and implicate *ADAMTS7* and *IL6R*, and related genes.

## Results

Our primary study population included 450,899 participants in the UK Biobank, who were mean (standard deviation) age 56.6 (8.0) years, 53.9% female, 45.0% ever smokers, and 83.6% European ancestry. At baseline, the prevalences of CAD, pneumonia, hypertension, hypercholesterolemia, and type 2 diabetes were 3.1%, 2.1%, 29.3%, 15.1%, and 2.5% respectively. Over 11.1-year median follow up, the number of incident cases of CAD, pneumonia, and type 2 diabetes were 13,374 (3.1%), 18,574 (4.2%), and 23,007 (5.2%), respectively (**Table 1**). The workflow for our study is depicted in **Supplemental Figure 1**.

**Table 1.** Characteristics of the study population in the UK Biobank (N= 450,899).

| **Metric** | **Value<sup>b</sup>** |
| --- | --- |
| Age (years)<sup>c</sup> | 56.6 (8.0) |
| Male | 207,968 (46.1%) |
| European ancestry | 376,785 (83.6%) |
| Ever smoked | 203,026 (45.0%) |
| Coronary artery disease | 13,797 (3.1%) |
| Pneumonia | 9,687 (2.1%) |
| Hypertension | 132,186 (29.3%) |
| Hypercholesterolemia | 68,089 (15.1%) |
| Type 2 diabetes | 11,136 (2.5%) |
| **Events During Follow-Up:** |  |
| Incident coronary artery disease | 13,374 (3.1%) |
| Incident pneumonia | 18,574 (4.2%) |
| Incident type 2 diabetes | 23,007 (5.2%) |

The study population was restricted to unrelated individuals in the UK biobank with unrelatedness defined as less than 3rd degree relatedness. Metrics are represented as mean (standard deviation) for continuous variables and % (n) for categorical variables. Clinical conditions are those occurring prior to enrollment unless noted as incident.

### Genome-wide pleiotropy search

With applied the PLeiotropic Analysis under COmposite null hypothesis (PLACO) to summary statistics for CAD from CARDIoGRAMplusC4D 2015^11^ and for pneumonia from FinnGen v4. A total of 115 SNPs with significant genetic pleiotropy between CAD and pneumonia defined as *p*_PLACO_<5 × 10^−8^ were identified. Among them, 88 also demonstrated evidence of association (*p*<0.05) with CAD and pneumonia risk in an independent dataset, which was a meta-analysis of the UK Biobank and Mass General Brigham Biobank. The CAD and pneumonia effects of all replicated SNPs were of opposite directions. These SNPs mapped to 2 distinct loci, and the annotated closest genes were *ADAMTS7* and *IL6R* (**Figure 1 and Supplemental Table 1**).

**Figure 1.**
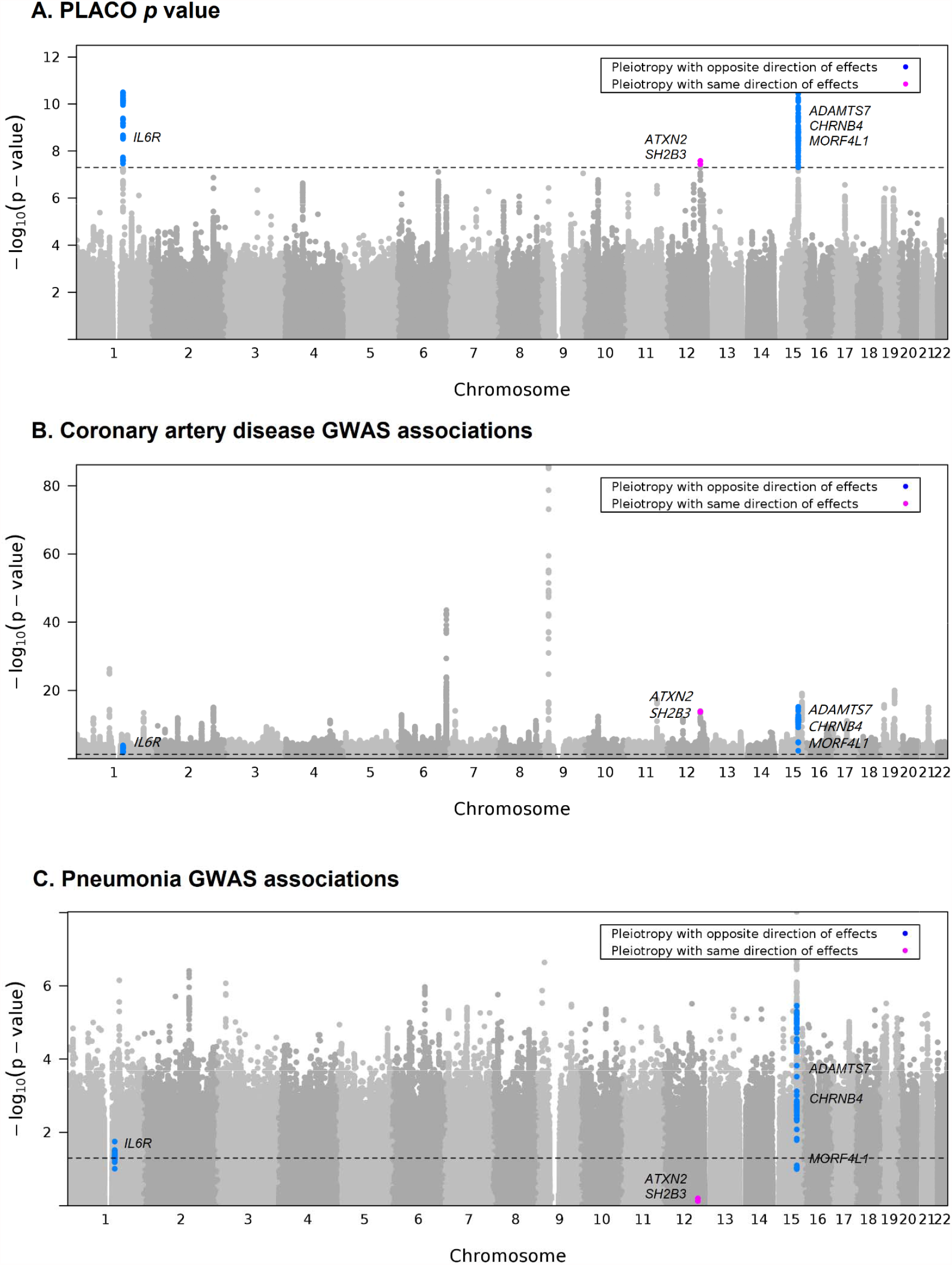
Manhattan plot of the PLACO p values (A) and results from the GWAS of CAD (B) and pneumonia (C). Genetic pleiotropy is defined as PLACO p values below genome-wide significance (5.0 × 10^−8^) with input GWAS of CAD from CARDIoGRAMplusC4D and that of pneumonia from FinnGen. GWAS results of CAD and pneumonia are from meta-analysis of UK Biobank and Mass General Brigham Biobank. 115 SNPs with significant evidence of genetic pleiotropy between CAD and pneumonia are highlighted: 113 SNPs with opposite direction of effects on CAD and pneumonia (nearest genes: *IL6R, CHRNB4, ADAMTS7, MORF4L1*) are highlighted in blue; 2 SNPs with same direction of effects (nearest genes: *SH2B3, ATXN2*) are highlighted in purple. The black horizontal dashed line in (A) corresponds to genome-wide significance (5E-8) and that in (B) and (C) corresponds to nominal significance (0.05). CAD: coronary artery disease, GWAS: genome-wide association study.

### Causal relations examination

We used eQTL data from the GTEx v8 to perform Mendelian randomization (MR) to assess the tissue specific effects of increased *ADAMTS7* and *IL6R* expression on CAD and pneumonia risk^12^. Based on known biology, the genetic effects of tissue-specific expression of *ADAMTS7* were assessed in aorta, tibial artery, and left ventricle and those of *IL6R* were assessed in tibial artery and whole blood. Across all those tissues, we observed that genetically increased *ADAMTS7* expression consistently correlated with significantly decreased risk for CAD and significantly increased risk for pneumonia, and genetically increased *IL6R* expression correlated with significantly increased risk for CAD and significantly decreased risk for pneumonia. Using inverse variance–weighted fixed-effects (IVW-FE) method, one standard deviation increase in the normalized gene expression of *ADAMTS7* was genetically associated with 0.85 odds (95% CI: 0.83-0.88; *p*=9.1 × 10^−28^) for CAD and 1.07 odds (95% CI: 1.03-1.11; *p*: 0.001) for pneumonia in tibial artery, and one standard deviation increase in genetic *IL6R* expression was associated with 1.18 odds (95% CI: 1.13-1.24; *p*=1.1 × 10^−11^) for CAD and 0.86 odds (95% CI: 0.80-0.93; *p*: 0.0001) for pneumonia in the same tissue (**Figure 2**). Sensitivity analyses using other MR methods, including summary-data-based MR (SMR), MR-Egger, weighted median, weighted mode, and MR RAPS, showed consistent results (**Supplemental Figure 2 and Supplemental Table 2**).

**Figure 2.**
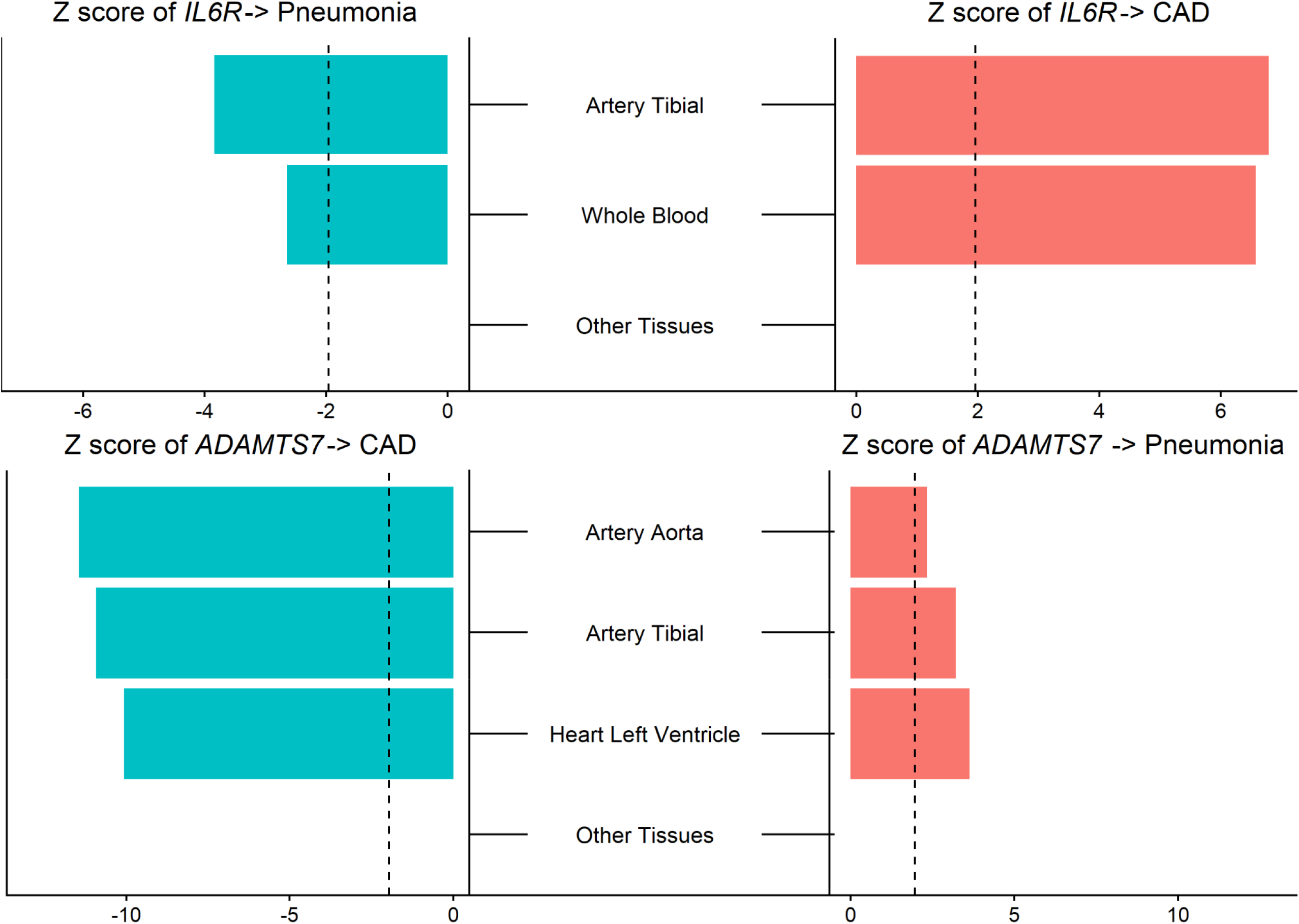
Tissue specific causal effects of the expression of *ADAMTS7* and *IL6R* gene on CAD and pneumonia. Mendelian randomization with inverse-variance weighted methods were used for this analysis. eQTL data were from GTEx v8. Relevant tissues included whole blood, spleen, small intestine terminal ileum, lung, liver, heart left ventricle, heart atrial appendage, artery tibial, artery coronary, artery aorta, adrenal gland, adipose visceral omentum, and adipose subcutaneous. Red bars indicate positive causal relations. Green bars indicate negative causal relations. The methods use was Black dashed lines indicate Z scores of 1.96, corresponding to p=0.05. CAD: coronary artery disease

Among the top ten genes whose corresponding proteins closely interact with ADAMTS7 protein according to STRING (**Supplemental Figure 3A**), three (*ADAMTS13, ADAMTSL1*, and *ADAMTSL3*) had valid eQTL gene expression for tibial artery in GTEx. None of the eQTL instruments for the three genes showed significant associations with either CAD or pneumonia (**Figure 3A**). As the expression of *IL6R* is well-recognized to be influenced by the concentrations of IL1B and NLRP3, we seeded STRING with *IL6R, IL1B*, and *NLRP3* for interacting genes in this pathway. Combining the top ten genes whose corresponded proteins closely interact with each of IL6R, IL1B, and NLRP3 proteins resulted in a total of 28 distinct genes, among which 24 had valid eQTL instruments for their expression concentrations in whole blood of data from the eQTLGen consortium, including *IL6, IL6ST, JAK1, JAK2, JAK3, STAT1, SAT3, STAT6, TYK2, IL1R1, IL1R2, IL1RAP, IL4, IL10, IL18, TNF, AIM2, DHX33, CASP1, CARD5, CARD8, NEK7, NLRC4*, and *TXNIP*. We tested *IL6R, IL1B*, and *NLRP3* together with the 24 genes, and found three of them had significant effects on both CAD and pneumonia: *IL6R, NLRP3*, and *DHX33*. Consistent with the results using tissue specific eQTL data from the GTEx, increased *IL6R* expression showed increased risk for CAD and decreased risk for pneumonia (*p*_CAD_=3.8 × 10^−4^; *p*_pneumonia_=0.03); the expression of *NLRP3* also demonstrated opposing directions of effects but with increased expression leading to decreased CAD risk and increased pneumonia risk (*p*_CAD_=0.009; *p*_pneumonia_=0.004). ; decreased *DHX33* gene expression was associated with decreased risk for both CAD and pneumonia (*p*_CAD_=0.009; *p*_pneumonia_=0.008) (**Figure 3B-D**). Our sensitivity analysis with using only cis-eQTL data for selecting instruments or using multiple MR methods yielded consistent results (**Supplemental Figure 4-5 and Supplemental Table 3**). When limiting to cis-eQTLs only, similar to *NLRP3*, decreased expression of the related *AIM2* inflammasome was associated with decreased CAD risk and increased pneumonia risk (**Supplementary Figure 4C**).

**Figure 3.**
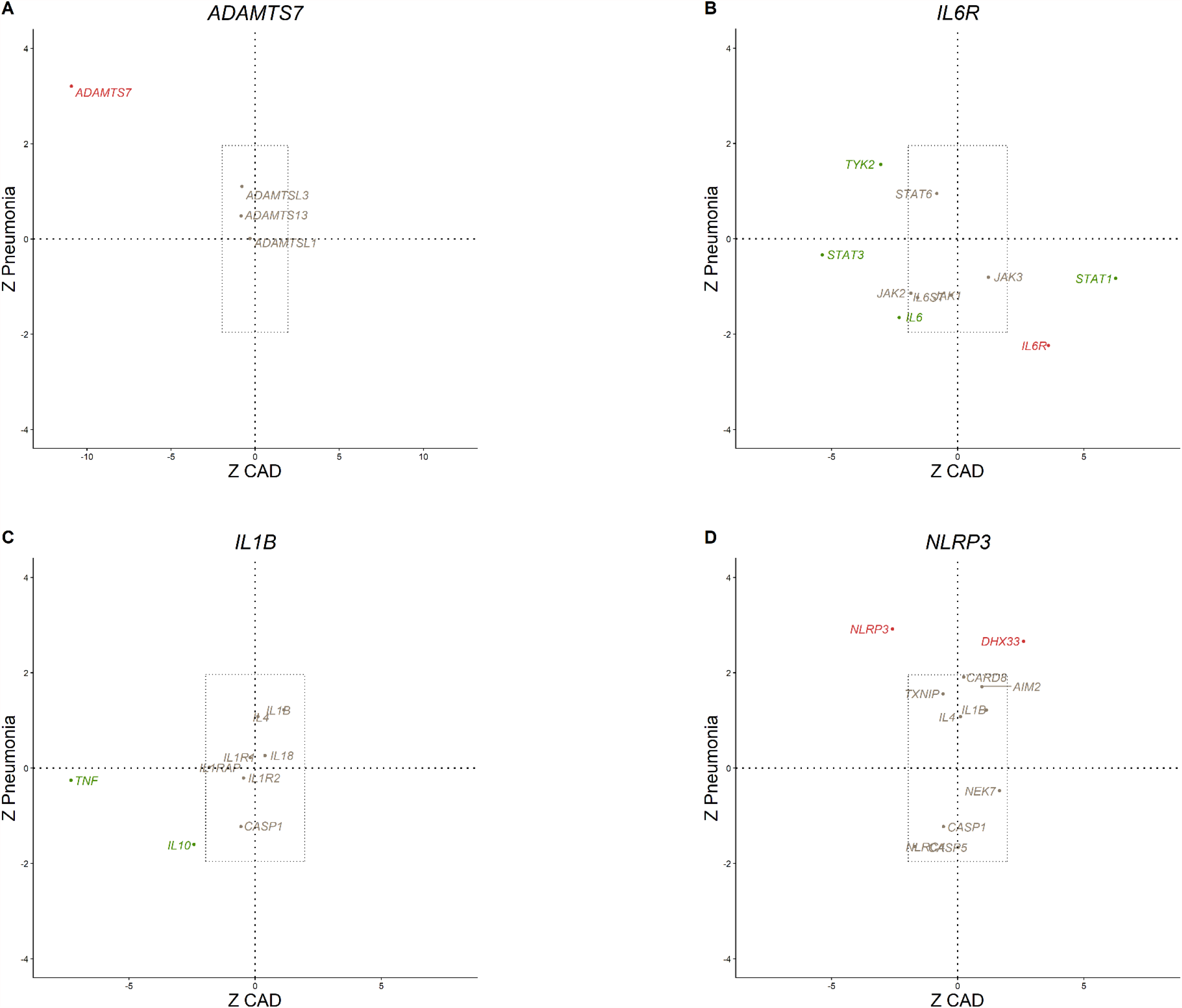
Causal effects of the expression of genes interacting with (A) *ADAMTS7*, (B) *IL6R*, (C) *IL1B*, and (D) *NLRP3* on CAD and pneumonia. eQTL data derived from tibial artery tissue for *ADAMTS7* related genes were from the GTEx v8. eQTL data derived from whole blood for *IL6R, IL1B, and NLRP3* related genes were from the eQTLGen consortium. Mendelian randomization with inverse-variance weighted methods were used for this analysis. Red dots indicate significant causal relations with both CAD and pneumonia. Green dots indicate significant causal relations with either CAD or pneumonia. Grey dots indicate significant causal relations with neither CAD nor pneumonia. Black dashed lines indicate Z scores of 1.96, corresponding to p=0.05. CAD: coronary artery disease, eQTL: expression quantitative trait loci.

### Gene-environment interaction examination

Based on existing literature, we identified *ADAMTS7* p.Ser214Pro (rs3825807, allele frequency=43.8%) as the genetic proxy for the function inhibition for *ADAMTS7*^13, 14^ and *IL6R* p.Asp358Ala (rs2228145, allele frequency=40.1%) for *IL6R*^8, 15, 16^, both of which were also identified as pleiotropic SNPs by PLACO. In the UK Biobank, among participants who never smoked, the presence of *ADAMTS7* p.Ser214Pro was associated with a significantly reduced risk of incident CAD (HR: 0.89; 95% CI: 0.84–0.93; *p*=1.1 × 10^−5^) for never smokers but not those who ever smoked (HR: 0.96; 95% CI: 0.92–1.01; *p*=0.14), revealing a statistical interaction between smoking status and *ADAMTS7* p.Ser214Pro (*p*_interaction=_0.01). In contrast, the presence of *ADAMTS7* p.Ser214Pro was associated with a significantly increased risk of pneumonia only among those who ever smoked (HR: 1.08; 95% CI: 1.03–1.12; *p*=4.4 × 10^−4^) but not those who never smoked (HR: 0.96; 95% CI: 0.92-1.01; *p*=0.15; *p*_interaction=_2.8 × 10^−4^) (**Table 2**). We did not observed statistically significant interactions between *IL6R* p.Asp358Ala and smoking status on either CAD or pneumonia.

**Table 2.** Coronary artery disease and pneumonia event incidence stratified by smoking status and genetically determined *ADAMTS7* level.

|  | Coronary artery disease<sup>1,2</sup> |  |  |  |  | Pneumonia<sup>1,3</sup> |  |  |  |  |
| --- | --- | --- | --- | --- | --- | --- | --- | --- | --- | --- |
|  | At risk | Incident events | P value | Hazard ratio | P for interaction | At risk | Incident events | P value | Hazard ratio | P for interaction |
| Non-smoker | 77,938 | 2,023 | Ref | 1.000 | 0.01 | 78,090 | 2,392 | Ref | 1.000 | $2.8 \times 10^{-4}$ |
| Non-smoker, *ADAMTS7* p.Ser214Pro | 165,159 | 3,875 | $1.1 \times 10^{-5}$ | 0.885 | | 165,137 | 4,899 | 0.15 | 0.964 | |
| Ever smoked | 60,835 | 2377 | Ref | 1.000 |  | 62,282 | 3358 | Ref | 1.000 |  |
| Ever smoked, *ADAMTS7* p.Ser214Pro | 133,082 | 5099 | 0.14 | 0.964 | | 135,615 | 7925 | $4.4 \times 10^{-4}$ | 1.075 | |

1. Model adjusted for age, age<sup>2</sup>, sex, race, first 10 principal components of ancestry, diagnosis of type 2 diabetes mellitus, and genotyping array.
2. Study population included 437,014 unrelated individuals without prevalent coronary artery disease (CAD) in the UK Biobank with unrelatedness defined as less than 3rd degree relatedness. CAD was defined as self-reported, hospitalization with, or death due to myocardial infarction; or hospitalization with OPCS4 (Office of Population Censuses and Surveys) codes for coronary artery bypass grafting (K40, K41, and K45) or coronary angioplasty with or without stenting (K49, K50.2, and K75).
3. Study population included 441,124 unrelated individuals without prevalent pneumonia in the UK Biobank with unrelatedness defined as less than 3rd degree relatedness. Pneumonia was defined as self-reported, hospitalization with, or death due to pneumonia.

## Discussion

Using a genome-wide pleiotropy scan, we demonstrated a genetic immune-inflammatory axis of CAD linked to pneumonia implicating *ADAMTS7* and *IL6R*, and related genes. For both genes as well as *NLRP3*, genetic analyses were consistent with opposing causal effects on CAD and pneumonia. We also found significant interactions between the expression of *ADAMTS7* and smoking status on incident CAD as well as pneumonia risk, indicating a cardioprotective effect without heighted pneumonia risk among nonsmokers. Our study provides a framework leveraging genetic pleiotropy to yield new biological and therapeutic insights.

Our approach uses a hypothesis-driven approach to examine immunity and CAD through concomitant analysis of pneumonia, a common co-morbid infectious disease. Observational epidemiology and translational research efforts have led to significant progress in improving the understanding of the pathophysiology underlying CAD during the past decades^17, 18^. However, despite effective treatment for traditional risk factors such as lipids and hypertension, CAD remains the leading causes of mortality and DALYs among population above age 50^1^. Recent pandemics of influenza, COVID-19, and other respiratory infections brought to the forefront their complex interplay with atherosclerosis, highlighting the potential separate risk of immune-inflammation on CAD^2, 3^. Probing the genetic correlation between CAD and pneumonia revealed the involvement of the immune-inflammatory genes *ADAMTS7* and *IL6R*.

Our results may have important implications for the prevention of CAD. First, smoking status may provide important information towards precision medicine immunomodulatory approaches for CAD. *ADAMTS7* belongs to the ADAMTS family of secreted metalloproteinases characterized with at least one thrombospondin type I repeat domains^19, 20^. Genome wide association studies (GWAS) previously revealed and independently replicated associations between multiple genetic variants at the *ADAMTS7* locus and susceptibility to CAD^21, 22, 23^, among which rs3825807 represents an A to G coding SNP, resulting in replacement of Ser214 by Pro thus affecting protein function^23^. *ADAMTS7* has also been validated as the causal gene by both *in vitro* and *in vivo* experiments^24, 25^. Expression of *ADAMTS7* in vascular smooth muscle cells (VSMCs) rises with inflammation but not with hyperlipidemia^25^. ADAMTS7 concentrations in human atherosclerotic plaque link to histological characteristics of plaque instability^26^. Our study demonstrated that *ADAMTS7* participates in CAD and pneumonia but in opposing directions. Prior analyses indicated that *ADAMTS7* is an essential gene for influenza A viral replication^27^. Mice deficient for *ADAMTS7* may also have greater influenza viral replication. Moreover, not only did we independently observe that *ADAMTS7* p.Ser214Pro confers CAD protection in never-smokers and not those who ever smoked^28^, we also found that *ADAMTS7* p.Ser214Pro significantly increased risk of pneumonia only among ever-smokers but not never-smokers. These findings indicate that therapeutic targeting of ADAMTS7 may be most effective in never-smokers – i.e., greater decrease in CAD risk without increased pneumonia risk.

Second, our approach aims to genetically prioritize therapeutic targets for CAD in the NLRP3/IL1B/IL6 pathway. Previously, the common *IL6R* p.Asp358Ala was shown to be associated with reduced cardiovascular disease risk^16, 29^. The Canakinumab Anti-inflammatory Thrombosis Outcomes Study (CANTOS) showed that, compared to placebo, patients with CAD and elevated high sensitivity C-reactive protein concentrations had reduced cardiovascular disease risk with canakinumab, a monoclonal antibody targeting IL1B^10^. Furthermore, the therapeutic effect was closely tied to the extent of IL6 concentration reduction^30^. Consistent with our human genetic observations, canakinumab versus placebo yielded a higher incidence of fatal infection in CANTOS^10^. Currently, there are multiple inhibitors of IL6 and NLRP3 that are in clinical development^31, 32^. We now provide human genetic validation for NLRP3 inhibition and CAD risk. However, our analyses indicate the possibility of increased infection risk as observed in CANTOS. While our germline genetic analyses model organism-wide perturbations, pharmacologic specificity may overcome such anticipated on-target adverse effects. In secondary analyses of interacting proteins, we observed that decreased *DHX33* gene expression was associated with decreased risk for both CAD and pneumonia. Therefore, alternate related targets may more optimally modulate immunity for both CAD and infectious disease.

The current study is strengthened by our robust design for genome-wide pleiotropy scan and causal relationship examinations: (1) in addition to having statistically significant evidence on pleiotropy, defined as *p*_PLACO_<5 × 10^−8^, only SNPs whose associations with CAD and pneumonia replicated in independent cohorts were carried forward for further steps; (2) leveraging eQTL data from both GTEx and eQTLGen, our causal inference results of *IL6R* and *ADAMTS7* on CAD and pneumonia are consistent across tissues and data resources; (3) sensitivity analysis with applying multiple MR methods on both cis- and trans-, and cis-only eQTL data also demonstrate the robustness of our findings. Our study also has limitations. (1) Due to the unavailability of valid instruments for measurement of *ADAMTS7* in whole blood, the causal inferences for *ADAMTS7* and its interacting genes were conducted using eQTL data of tibial artery from GTEx, which has smaller sample size and only cis-eQTL data versus the *IL6R* related analyses which used data from the eQTLGen consortium. (2) Our gene-smoking interaction exploration dichotomized smoking status to ever-smoker and never-smoker without examining for the potential dose-response of smoking on disease risk due to the large number of missingness of corresponding variables in the UK Biobank. Heterogeneity among ever-smokers may exist. (3) While our human genetic study provides strong causal inference, perturbation in controlled model systems and prospective human randomized controlled trials are necessary to definitively establish causality.

In conclusion, leveraging novel statistical methods, eQTL data from multiple tissues and resources, together with large-scale population cohort, we demonstrated genetic immune-inflammatory axes of CAD linked to respiratory infections implicate *ADAMTS7* and *IL6R*, and related genes.

## Methods

### Study population

The UK Biobank is a prospective cohort consists of approximately 500,000 adult participants recruited between 2006 and 2010 from 22 assessment centers across the United Kingdom^33^. Our analysis included 450,899 unrelated individuals in the UK Biobank. Samples were excluded for the following reasons: genotypic sex did not match reported sex, kinship was not inferred, putative sex chromosome aneuploidy, consent withdrawn, or excessive heterozygosity or missingness, based on centralized sample quality control performed by UK Biobank. Excluded related individuals were defined as one individual in each pair within the third degree of relatedness determined based on kinship coefficients centrally calculated by UK Biobank ^33^. Outcomes of this analysis were incident CAD and pneumonia: CAD was defined as self-reported, hospitalization with, or death due to myocardial infarction; or hospitalization with OPCS4 (Office of Population Censuses and Surveys) codes for coronary artery bypass grafting (K40, K41, and K45) or coronary angioplasty with or without stenting (K49, K50.2, and K75). Pneumonia was defined as self-reported, hospitalization with, or death due to pneumonia. Prevalent cases were removed when analyzing the corresponding incident outcomes.

Mass General Brigham Biobank is an ongoing hospital-based cohort study of patients across the Mass General Brigham (formerly Partners Healthcare) health system. It is enriched with longitudinal electronic medical records (EMR) data, genomic data, and electronic health and lifestyle survey data^34^. The present secondary analyses were approved by the Massachusetts General Hospital Institutional Review Board.

### Genome-wide pleiotropy search

We applied PLeiotropic Analysis under COmposite null hypothesis (PLACO) to conduct a genome wide search for SNPs influencing risks of both CAD and pneumonia. Briefly, PLACO is a novel statistical method for identifying pleiotropic loci between two traits through testing the composite null hypothesis that a locus is associated with zero or one of the traits. It only needs summary statistics as input, and the test statistic is formed as the product of the Z statistics of the SNP in each of the two studies that assumed to follow a mixture distribution^35^. Input genome-wide summary statistics for CAD were from CARDIoGRAMplusC4D (Coronary ARtery DIsease Genome wide Replication and Meta-analysis (CARDIoGRAM) plus The Coronary Artery Disease (C4D) Genetics) 2015^11^ and those for pneumonia were from FinnGen R4^36^. CARDIoGRAMplusC4D 2015 is a meta-analysis of GWAS studies including 60,801 CAD case and 123,504 control participants with mainly European, South Asian, and East Asian ancestries^11^. The FinnGen Study is a Finnish, nationwide GWAS meta-analysis that linked with EMR data from national health registries. In R4, there were 20389 pneumonia cases defined as phenotype ICD10-J10 pneumonia and 156510 controls^36^. There were no overlap of study populations underlying the CAD and pneumonia GWAS. We removed SNPs with *Z*^2^ > 80 as extremely large significance may disproportionately influence the analysis^35, 37^. For pleiotropic SNPs with statistically significant evidence of genome-wide pleiotropy, defined as P_PLACO_ < 5 × 10^−8^, we independently assessed for nominal association with both CAD and pneumonia in a meta-analysis of UK Biobank^38^ and Mass General Brigham Biobank. We clumped variants in ±500 Kb radius and linkage disequilibrium (LD) threshold of *r*^2^>0.2 into a single genetic locus using FUMA (SNP2GENE function, v1.3.6a). The gene annotations for all loci are based on proximity to the most significant/lead SNPs as mapped by FUMA^39^.

### Genetic relations examination

We evaluated the genetic relations of tissue-specific expression of genes annotated to the pleiotropic SNPs on CAD and pneumonia using MR toward causal inference. eQTL data of 13 tissues relevant to CAD and pneumonia, i.e., whole blood, spleen, small intestine terminal ileum, lung, liver, heart left ventricle, heart atrial appendage, artery tibial, artery coronary, artery aorta, adrenal gland, adipose visceral omentum, and adipose subcutaneous, was obtained from the GTEx project (v8; https://gtexportal.org/home/). These analyses were limited to cis-eQTL, which were defined as variants within 1 Mb of the transcription start site (TSS) of each gene, given data availability^12^. Summary statistics of CAD were from meta-analysis of CARDIoGRAMplusC4D and UK Biobank^40^ and that of pneumonia were from meta-analysis of FinnGen R4^36^ and UK Biobank^38^.

We extended the causal relation evaluations for both identified pleiotropic genes to their interacting genes identified through STRING, a protein-protein interaction database, (v11; https://string-db.org/) using same MR approaches. For index pleiotropic gene(s) that are expressed in whole blood, we used eQTL data derived from whole blood in >31,000 individuals from the eQTLGen consortium (http://www.eqtlgen.org) for evaluating their and their interacting genes’ causal effects on CAD and pneumonia. As the eQTLGen consortium provided summary statistics of both cis- and trans-eQTL results, we included both cis- and trans-eQTL for primary analysis to maximize power and because recent studies showed that only a modest fraction of trait-heritability can be explained by cis-mediated bulk gene-expressions^41^. We also conducted sensitivity analysis with using cis-eQTL results only to address potential pleiotropy. As the eQTLGen consortium only included eQTL data derived from whole blood, for other index pleiotropic gene(s) and their interacting genes not highly expressed in whole blood, we used eQTL data of tibial artery, due to similarity to coronary artery, from the GTEx project. As the GTEx project only included cis-eQTL data, we used cis-eQTLs for primary analysis.

For both sets of causal relation analyses, we selected instruments from each dataset using independent (r^2^ < 0.05) SNPs that met genome-wide significance (P < 5 × 10^−8^). We applied two-sample MR to estimate the causal relationship between eQTLs and CVD and pneumonia using the MR-Base R package (version 0.5.5)^42^. For genes that were instrumented with a single SNP, Wald ratio MR method was used; for those with more than one valid instrument, we conducted MR analyses using inverse variance–weighted fixed-effects (IVW-FE) as the primary method^43^, as well as summary-data-based MR (SMR) method which is less prone to false-positive findings and other alternative MR methods (i.e., MR-Egger, weighted median, weighted mode, or MR RAPS) that may be more powerful under various model assumptions for sensitivity analyses^44, 45, 46, 47, 48^. As the use of SMR requires eQTL data in BESD format and provided GTEx v7 eQTL data in that format, we used GTEx v7 eQTL data when using SMR. A reference panel of European individuals from the phase 3 1000 Genomes project was used to compute linkage disequilibrium (LD) estimation for all analyses^49^.

### Gene-environment interaction examination in human

Based on existing literature, we identified one common (allele frequency > 30%) disruptive coding mutations of each pleiotropic gene as a genetic proxy for the function inhibition of that gene – *ADAMTS7* p.Ser214Pro and *IL6R* p.Asp358Ala. We performed a prospective, time-to-event analysis for the incident CAD and pneumonia outcomes in the UK Biobank stratified by the status of the instrument for each pleiotropic gene. Cox proportional hazard models were adjusted for age, age^2^, sex, race, first 10 genetic principal components (PCs), diagnosis of type 2 diabetes mellitus, and genotyping array. To test for proportional hazards assumption for the genetic instruments, we examined whether their corresponding scaled Schoenfeld residuals were independent of follow-up time and found no statistically significant evidence suggesting violation of the assumption. Race was determined by self-reported ancestry and followed by outlier detection based on genetic PCs centrally calculated by UK Biobank. Since smoking represents a key shared risk factor for both CAD and pneumonia, we evaluated the interaction between smoking status and each instrument on the primary outcomes. Analyses used R version 3.6.2 software (The R Foundation), two-tailed P-values, as well as statistical significance level of P < 0.05 except for the identification of genome-wide signals, which was P < 5 × 10^−8^.

## Data Availability

UKB individual-level data are available for request by application (https://www.ukbiobank.ac.uk). Individual-level MGBB data are available from https://personalizedmedicine.partners.org/Biobank/Default.aspx, but restrictions apply to the availability of these data. All the summary-level data used are available for download at the public repositories.

## Notes

### Competing Interest Statement

P.L. is an unpaid consultant to, or involved in clinical trials for, Amgen, AstraZeneca, Baim Institute, Beren Therapeutics, Esperion Therapeutics, Genentech, Kancera, Kowa Pharmaceuticals, Medimmune, Merck, Norvo Nordisk, Novartis, Pfizer and Sanofi-Regeneron. P.L. is a member of scientific advisory board for Amgen, Caristo, Cartesian, Corvidia Therapeutics, CSL Behring, DalCor Pharmaceuticals, Dewpoint, Kowa Pharmaceuticals, Olatec Therapeutics, Medimmune, Novartis, PlaqueTec and XBiotech, Inc. P.L.'s laboratory has received research funding in the past 2 years from Novartis. P.L. is on the Board of Directors of XBiotech, and has a financial interest in Xbiotech, a company developing therapeutic human antibodies. P.L.'s interests were reviewed and are managed by Brigham and Women's Hospital and Partners HealthCare in accordance with their conflict-of-interest policies. P.N. reports investigator-initiated grants from Amgen, Apple, Boston Scientific, and AstraZeneca, personal fees from Apple, Blackstone Life Sciences, Foresite Labs, Genentech, and Novartis, and spousal employment at Vertex, all unrelated to the present work.

### Funding Statement

H.K.F. was funded by NIH grant DP5 OD024582 and by Eric and Wendy Schmidt. P.N. is supported by grants from the National Heart, Lung, and Blood Institute (R01HL142711, R01HL148050, R01HL151283, R01HL148565, R01HL135242, R01HL151152), Fondation Leducq (TNE-18CVD04), and Massachusetts General Hospital (Paul and Phyllis Fireman Endowed Chair in Vascular Medicine).

### Author Declarations

Use of the UK Biobank data was approved by the Massachusetts General Hospital Institutional Review Board (protocol 2013P001840) and facilitated through UK Biobank Applications 7089 and 21552. Mass General Brigham Biobank analyses were approved by the Massachusetts General Hospital Institutional Review Board (protocol 2020P000904).

## Reference

1. Vos T, et al. Global burden of 369 diseases and injuries in 204 countries and territories, 1990-2019: a systematic analysis for the Global Burden of Disease Study 2019. The Lancet 396, 1204–1222 (2020).

2. Nishiga M, Wang DW, Han Y, Lewis DB, Wu JC. COVID-19 and cardiovascular disease: from basic mechanisms to clinical perspectives. Nat Rev Cardiol 17, 543–558 (2020).

3. Siddiqi HK, Libby P, Ridker PM. COVID-19 - A vascular disease. Trends Cardiovasc Med 31, 1–5 (2021).

4. Corrales-Medina VF, Musher DM, Shachkina S, Chirinos JA. Acute pneumonia and the cardiovascular system. The Lancet 381, 496–505 (2013).

5. Phrommintikul A, Kuanprasert S, Wongcharoen W, Kanjanavanit R, Chaiwarith R, Sukonthasarn A. Influenza vaccination reduces cardiovascular events in patients with acute coronary syndrome. Eur Heart J 32, 1730–1735 (2011).

6. Natarajan P, Cannon CP. Myocardial infarction vaccine? Evidence supporting the influenza vaccine for secondary prevention. Eur Heart J 32, 1701–1703 (2011).

7. Libby P, et al. Inflammation, Immunity, and Infection in Atherothrombosis: JACC Review Topic of the Week. J Am Coll Cardiol 72, 2071–2081 (2018).

8. Swerdlow DI, et al. The interleukin-6 receptor as a target for prevention of coronary heart disease: a mendelian randomisation analysis. Lancet 379, 1214–1224 (2012).

9. Sarwar N, et al. Interleukin-6 receptor pathways in coronary heart disease: a collaborative meta-analysis of 82 studies. Lancet 379, 1205–1213 (2012).

10. Ridker PM, et al. Antiinflammatory Therapy with Canakinumab for Atherosclerotic Disease. New England Journal of Medicine 377, 1119–1131 (2017).

11. Nikpay M, et al. A comprehensive 1,000 Genomes-based genome-wide association meta-analysis of coronary artery disease. Nat Genet 47, 1121–1130 (2015).

12. The GTEx Consortium atlas of genetic regulatory effects across human tissues. Science 369, 1318–1330 (2020).

13. Pu X, et al. ADAMTS7 Cleavage and Vascular Smooth Muscle Cell Migration Is Affected by a Coronary-Artery-Disease-Associated Variant. The American Journal of Human Genetics 92, 366–374 (2013).

14. Chan K, et al. Genetic Variation at the ADAMTS7 Locus is Associated With Reduced Severity of Coronary Artery Disease. Journal of the American Heart Association 6, e006928 (2017).

15. Cai T, et al. Association of Interleukin 6 Receptor Variant With Cardiovascular Disease Effects of Interleukin 6 Receptor Blocking Therapy: A Phenome-Wide Association Study. JAMA Cardiology 3, 849–857 (2018).

16. Bick AG, et al. Genetic Interleukin 6 Signaling Deficiency Attenuates Cardiovascular Risk in Clonal Hematopoiesis. Circulation 141, 124–131 (2020).

17. Patel AP, Natarajan P. Completing the genetic spectrum influencing coronary artery disease: from germline to somatic variation. Cardiovasc Res 115, 830–843 (2019).

18. Crea F, Libby P. Acute Coronary Syndromes: The Way Forward From Mechanisms to Precision Treatment. Circulation 136, 1155–1166 (2017).

19. Apte SS. A disintegrin-like and metalloprotease (reprolysin type) with thrombospondin type 1 motifs: the ADAMTS family. Int J Biochem Cell Biol 36, 981–985 (2004).

20. Apte SS. A disintegrin-like and metalloprotease (reprolysin-type) with thrombospondin type 1 motif (ADAMTS) superfamily: functions and mechanisms. J Biol Chem 284, 31493–31497 (2009).

21. Reilly MP, et al. Identification of ADAMTS7 as a novel locus for coronary atherosclerosis and association of ABO with myocardial infarction in the presence of coronary atherosclerosis: two genome-wide association studies. Lancet 377, 383–392 (2011).

22. A genome-wide association study in Europeans and South Asians identifies five new loci for coronary artery disease. Nat Genet 43, 339–344 (2011).

23. Schunkert H, et al. Large-scale association analysis identifies 13 new susceptibility loci for coronary artery disease. Nat Genet 43, 333–338 (2011).

24. Bauer RC, et al. Knockout of Adamts7, a Novel Coronary Artery Disease Locus in Humans, Reduces Atherosclerosis in Mice. Circulation 131, 1202–1213 (2015).

25. Hanby HA, Zheng XL. Biochemistry and physiological functions of ADAMTS7 metalloprotease. Adv Biochem 1, 10.11648/j.ab.20130103.20130111 (2013).

26. Bengtsson E, et al. ADAMTS-7 is associated with a high-risk plaque phenotype in human atherosclerosis. Scientific Reports 7, 3753 (2017).

27. Meliopoulos VA, et al. MicroRNA Regulation of Human Protease Genes Essential for Influenza Virus Replication. PLOS ONE 7, e37169 (2012).

28. Saleheen D, et al. Loss of Cardioprotective Effects at the ADAMTS7 Locus as a Result of Gene-Smoking Interactions. Circulation 135, 2336–2353 (2017).

29. The interleukin-6 receptor as a target for prevention of coronary heart disease: a mendelian randomisation analysis. The Lancet 379, 1214–1224 (2012).

30. Ridker PM, et al. Modulation of the interleukin-6 signalling pathway and incidence rates of atherosclerotic events and all-cause mortality: analyses from the Canakinumab Anti-Inflammatory Thrombosis Outcomes Study (CANTOS). Eur Heart J 39, 3499–3507 (2018).

31. Mezzaroma E, Abbate A, Toldo S. NLRP3 Inflammasome Inhibitors in Cardiovascular Diseases. Molecules 26, (2021).

32. Ridker PM, et al. IL-6 inhibition with ziltivekimab in patients at high atherosclerotic risk (RESCUE): a double-blind, randomised, placebo-controlled, phase 2 trial. The Lancet 397, 2060–2069 (2021).

33. Bycroft C, et al. The UK Biobank resource with deep phenotyping and genomic data. Nature 562, 203–209 (2018).

34. Karlson EW, Boutin NT, Hoffnagle AG, Allen NL. Building the Partners HealthCare Biobank at Partners Personalized Medicine: Informed Consent, Return of Research Results, Recruitment Lessons and Operational Considerations. J Pers Med 6, (2016).

35. Ray D, Chatterjee N. A powerful method for pleiotropic analysis under composite null hypothesis identifies novel shared loci between Type 2 Diabetes and Prostate Cancer. PLOS Genetics 16, e1009218 (2020).

36. FinnGen. FinnGen: Documentation of R4 release.).

37. Bulik-Sullivan BK, et al. LD Score regression distinguishes confounding from polygenicity in genome-wide association studies. Nature Genetics 47, 291–295 (2015).

38. Zekavat SM, et al. Elevated Blood Pressure Increases Pneumonia Risk: Epidemiological Association and Mendelian Randomization in the UK Biobank. Med 2, 137-148.e134 (2021).

39. Watanabe K, Taskesen E, van Bochoven A, Posthuma D. Functional mapping and annotation of genetic associations with FUMA. Nat Commun 8, 1826 (2017).

40. Nelson CP, et al. Association analyses based on false discovery rate implicate new loci for coronary artery disease. Nat Genet 49, 1385–1391 (2017).

41. Yao DW, O’Connor LJ, Price AL, Gusev A. Quantifying genetic effects on disease mediated by assayed gene expression levels. Nat Genet 52, 626–633 (2020).

42. Hemani G, et al. The MR-Base platform supports systematic causal inference across the human phenome. Elife 7, (2018).

43. Burgess S, Butterworth A, Thompson SG. Mendelian randomization analysis with multiple genetic variants using summarized data. Genet Epidemiol 37, 658–665 (2013).

44. Wu Y, et al. Integrative analysis of omics summary data reveals putative mechanisms underlying complex traits. Nature Communications 9, 918 (2018).

45. Bowden J, Davey Smith G, Haycock PC, Burgess S. Consistent Estimation in Mendelian Randomization with Some Invalid Instruments Using a Weighted Median Estimator. Genet Epidemiol 40, 304–314 (2016).

46. Schmidt AF, Dudbridge F. Mendelian randomization with Egger pleiotropy correction and weakly informative Bayesian priors. International Journal of Epidemiology 47, 1217–1228 (2017).

47. Hartwig FP, Davey Smith G, Bowden J. Robust inference in summary data Mendelian randomization via the zero modal pleiotropy assumption. Int J Epidemiol 46, 1985–1998 (2017).

48. Zhao Q, Wang J, Hemani G, Bowden J, Small DS. Statistical inference in two-sample summary-data Mendelian randomization using robust adjusted profile score. arXiv 1801.09652, (2018).

49. McVean GA, et al. An integrated map of genetic variation from 1,092 human genomes. Nature 491, 56–65 (2012).

