## Supplemental material for "Genome-wide pleiotropy analysis of coronary artery disease and pneumonia identifies shared immune pathways"

GWAS of pneumonia from FinnGen R4

GWAS of CAD from CARDIoGRAMplusC4D 2015

115 SNPs on 6 nearest genes (*IL6R, CHRNB4, ADAMTS7, MORF4L1, SH2B3, ATXN2*) with significant evidence of genetic pleiotropy

88 SNPs present and reached nominal significant in the meta-analysis of GWAS for CAD and pneumonia from the UK Biobank and Mass General Brigham biobank

Clumped to 2 distinctive loci on *IL6R* and *ADAMTS7*

Examine causal relations of *IL6R* and *ADAMTS7* on CAD and pneumonia using Mendelian randomization using eQTL summary statistics of 13 relevant tissues from GTEX v8

Expand *IL6R* to 25 genes based on (1) *IL6R* signaling pathway and (2) protein-protein interaction from STRING and examine causal relations of *IL6R* and related genes on CAD and pneumonia using eQTL results of whole blood from eQTLGen consortium

Expand *ADAMTS7* to 4 genes based on protein-protein interaction from STRING and Examine causal relations of *ADAMTS7* and related genes on CAD and pneumonia using eQTL results of tibial artery from GTEX v8

Examine for interactions between *IL6R* p.Asp358Ala and smoking status on incident CAD and pneumonia risks in UK Biobank (N=450,899)

Examine for interactions between *ADAMTS7* p.Ser214Pro and smoking status on incident CAD and pneumonia risks in UK Biobank (N=450,899)

**Supplemental Figure 1. Study design and workflow.** Genome-wide pleiotropy analyses of CAD and pneumonia using summary statistics from CARDIoGRAMplusC4D 2015 (CAD) and FinnGen R4 (pneumonia) identified 115 SNPs with significant evidence of genetic pleiotropy, among which 88 were independently replicated in the UK Biobank and Mass General Brigham biobank and were clumped to 2 distinctive loci annotated to *IL6R* and *ADAMTS7*. Causal effects of *IL6R* and *ADAMTS7*, as well as their interacting genes, on CAD and pneumonia were examined using Mendelian randomization using eQTL summary statistics from the eQTLGen consortium and/or GTEX v8. Potential interactions between disruptive coding mutations of *IL6R* and *ADAMTS7* and smoking status on incident CAD and pneumonia risks were examined in UK Biobank.

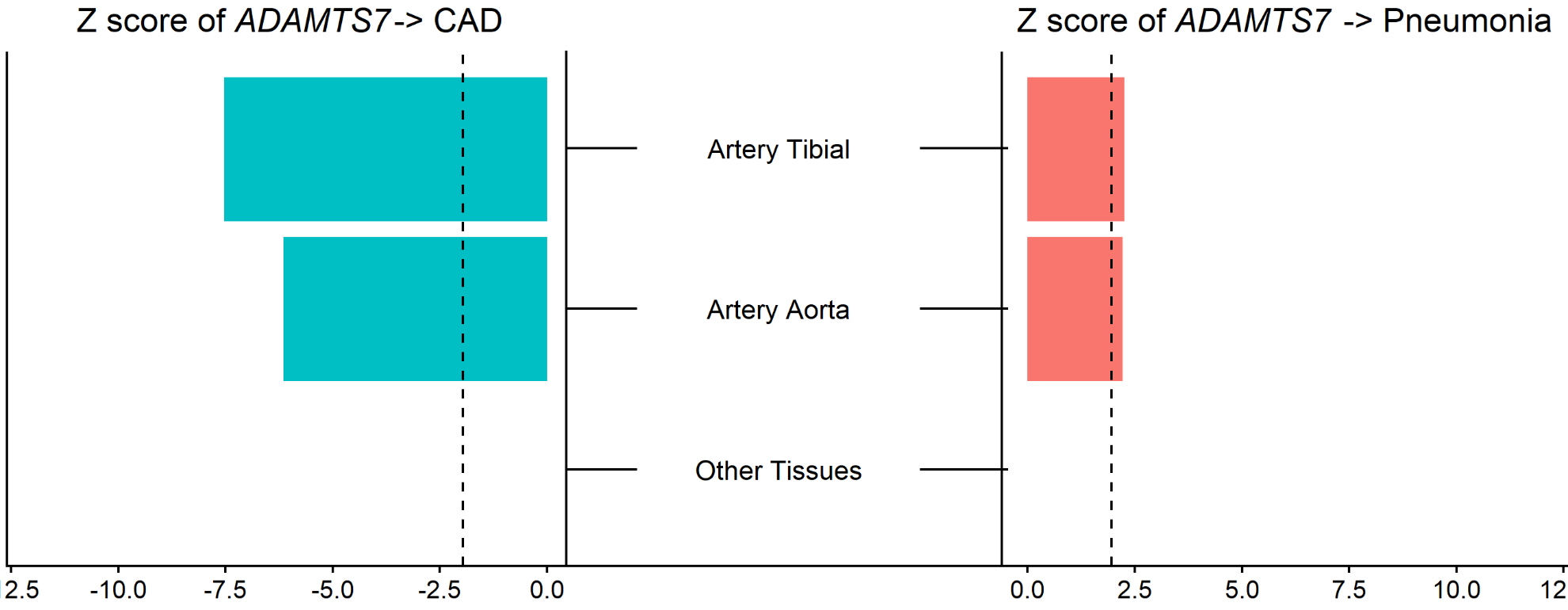

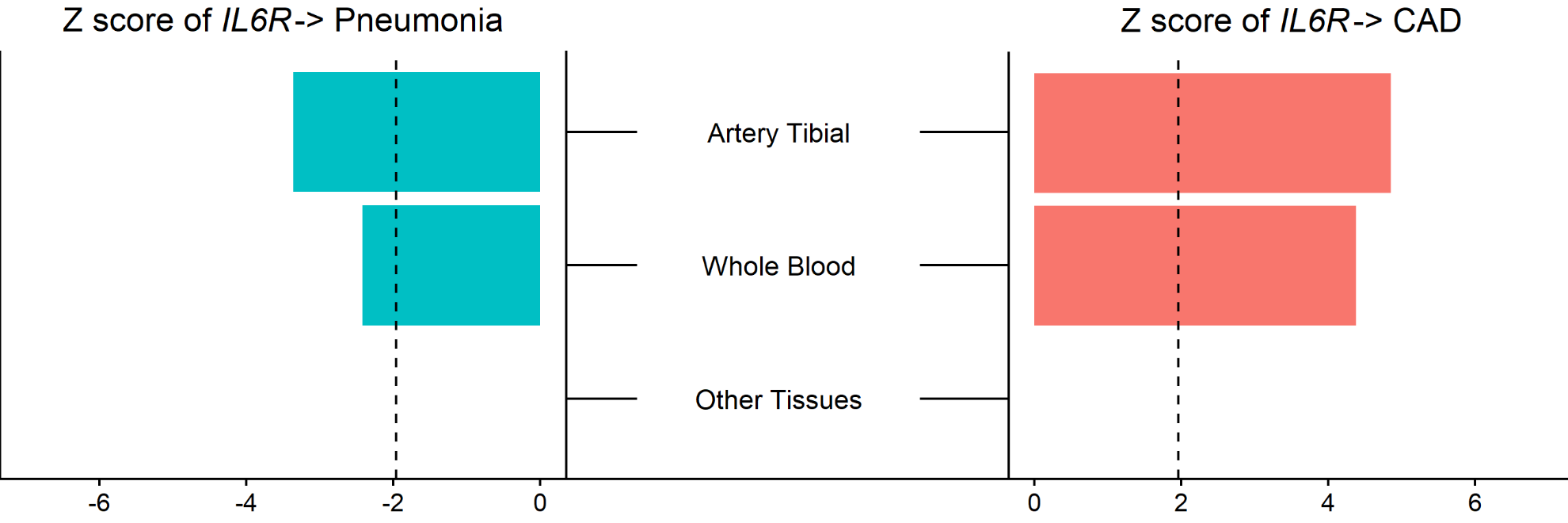

###### Supplemental Figure 2. Causal effects of the expression of *ADAMTS7* and *IL6R* gene on CAD and pneumonia using eQTL summary statistics across relevant tissues. Mendelian randomization with summary data–based Mendelian randomization (SMR) methods were used for this analysis. eQTL summary statistics were from GTEx v7. Relevant tissues included whole blood, spleen, small intestine terminal ileum, lung, liver, heart left ventricle, heart atrial appendage, artery tibial, artery coronary, artery aorta, adrenal gland, adipose visceral omentum, and adipose subcutaneous. Red bars indicate positive causal relations. Green bars indicate negative causal relations. Black dashed lines indicate Z scores of 1.96, corresponding to p=0.05. CAD: coronary artery disease

| A *ADAMTS7*  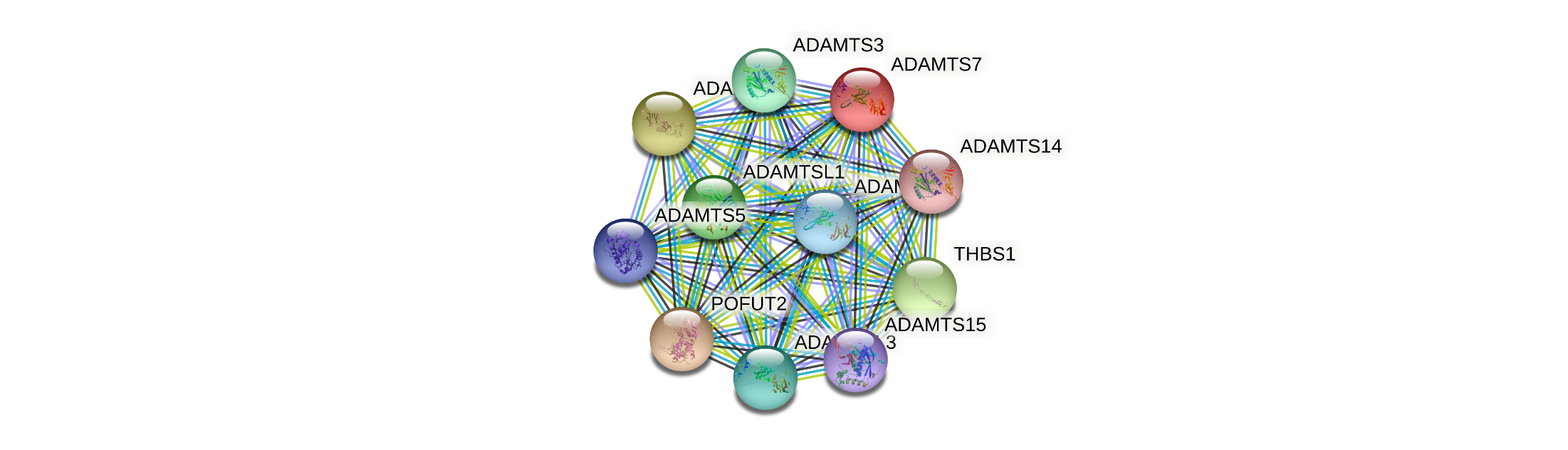 | B *IL6R*  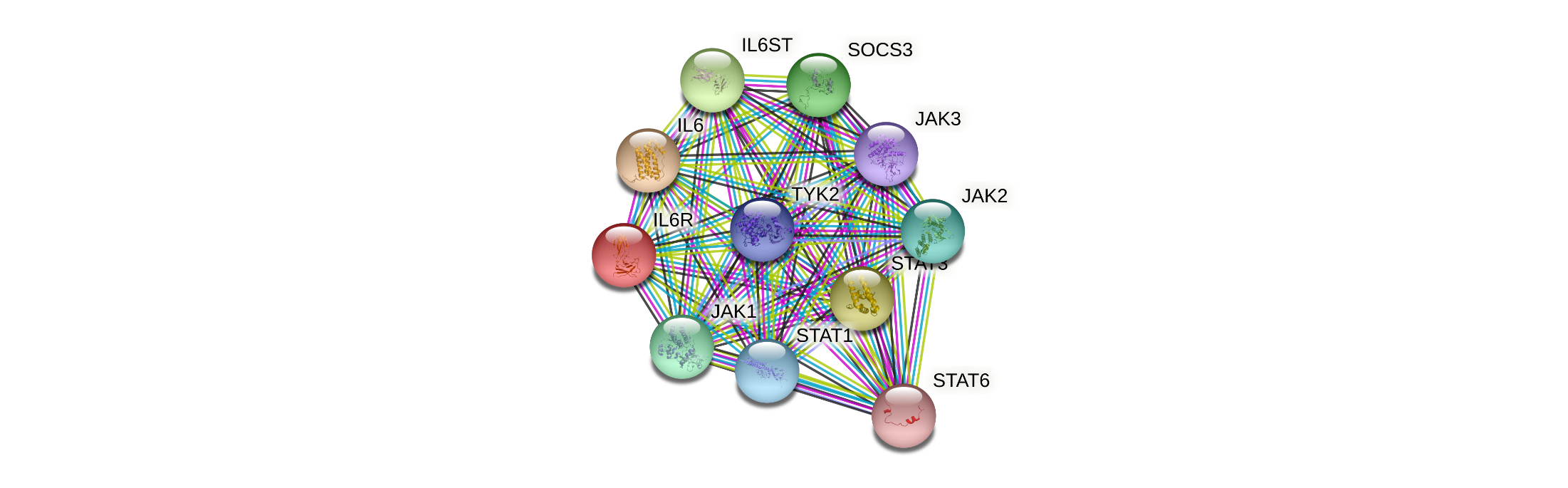 |
| --- | --- |
| C *IL1B*  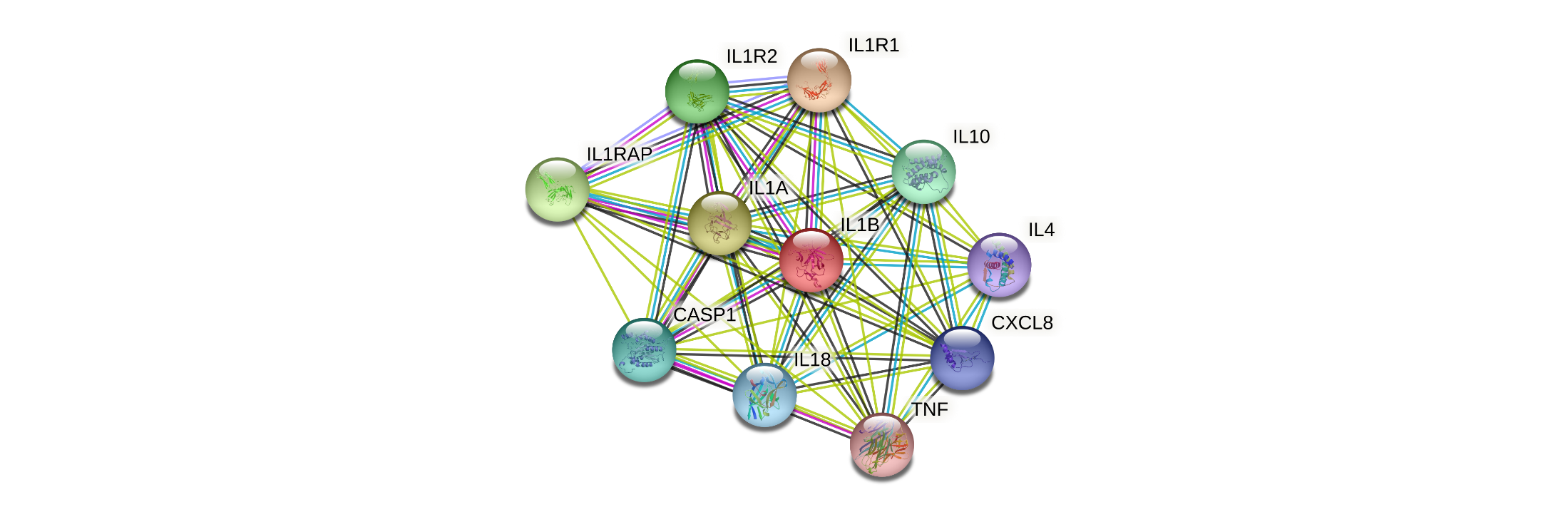 | D *NLRP3*  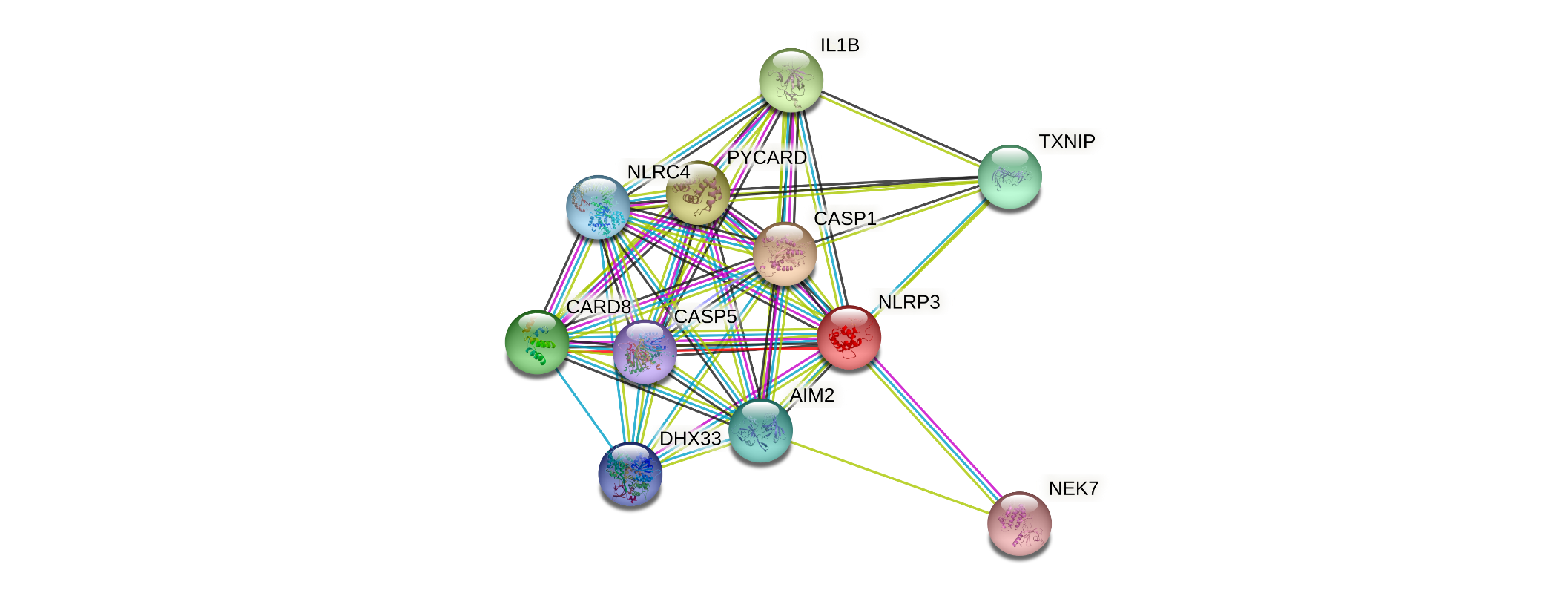 |

###### Supplemental Figure 3. Protein-protein interaction network of (A) ADAMTS7, (B) IL6R, (C) IL1B, and (D) NLRP3. Network structures were obtained from STRING database (v11; https://string-db.org/).

| 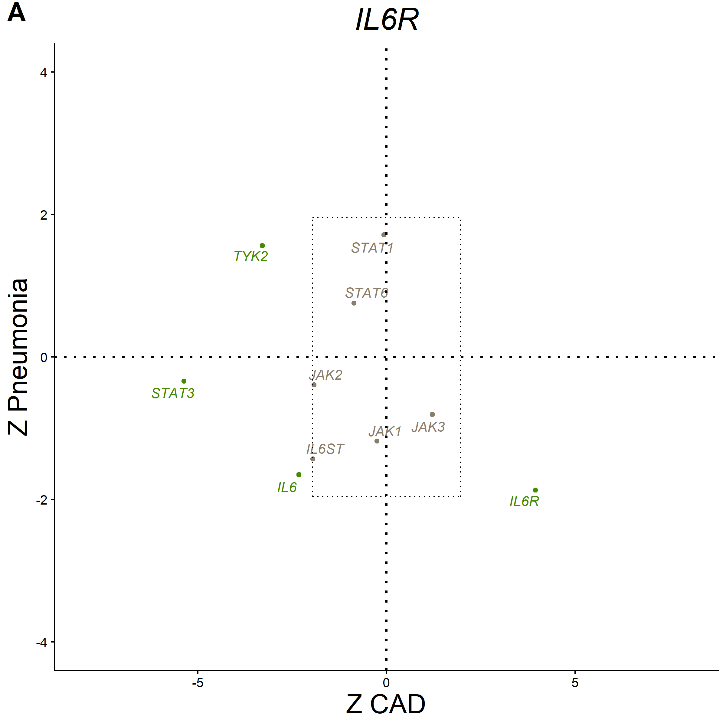 | 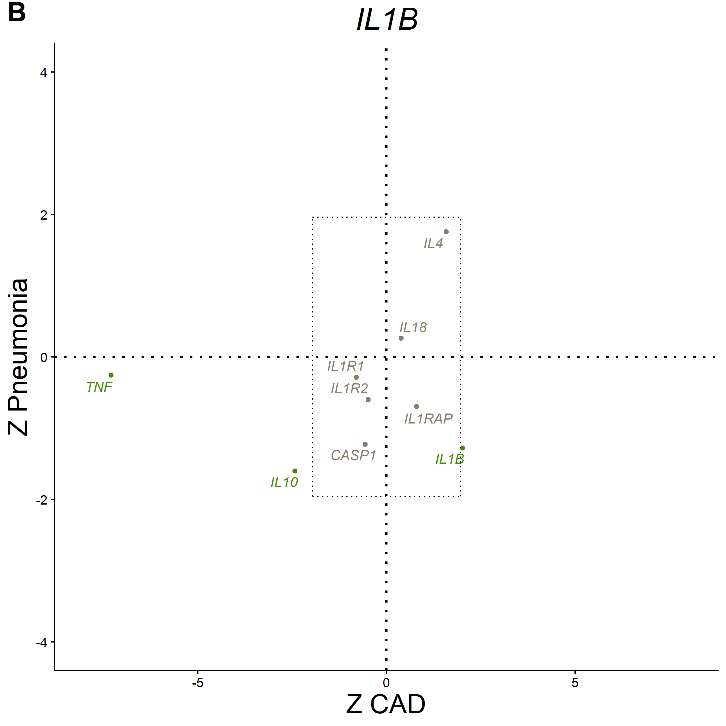 |
| --- | --- |
| 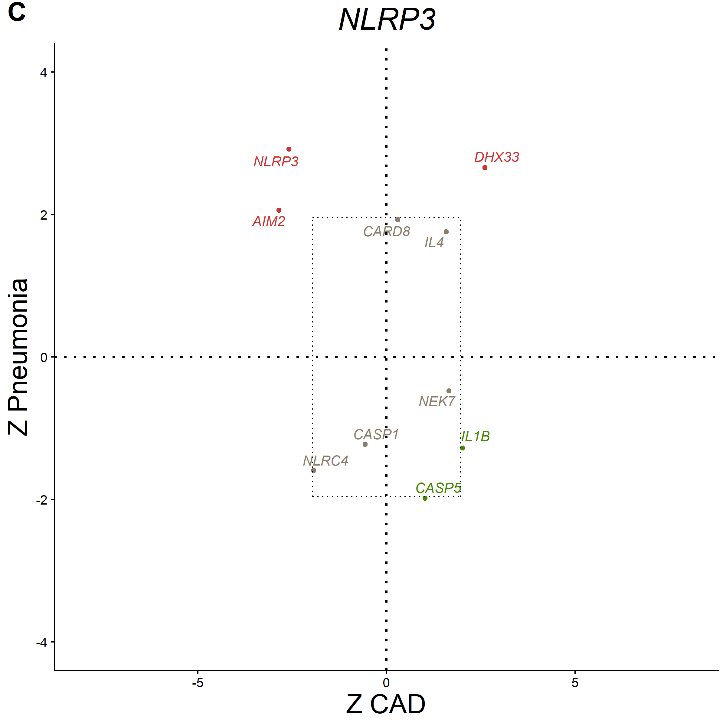 |  |

###### Supplemental Figure 4. Causal effects of the expression of genes interacting with (A) *IL6R*, (B) *IL1B*, and (C) *NLRP3* on CAD and pneumonia using cis-eQTL data. cis-eQTL summary statistics for *IL6R, IL1B, and NLRP3* related biomarkers were from whole blood of the eQTLGen consortium. Mendelian randomization with inverse-variance weighted methods were used for this analysis. Red dots indicate significant causal relations with both CAD and pneumonia. Green dots indicate significant causal relations with either CAD or pneumonia. Grey dots indicate significant causal relations with neither CAD nor pneumonia. Black dashed lines indicate Z scores of 1.96, corresponding to p=0.05. CAD: coronary artery disease, eQTL: expression quantitative trait loci.

| 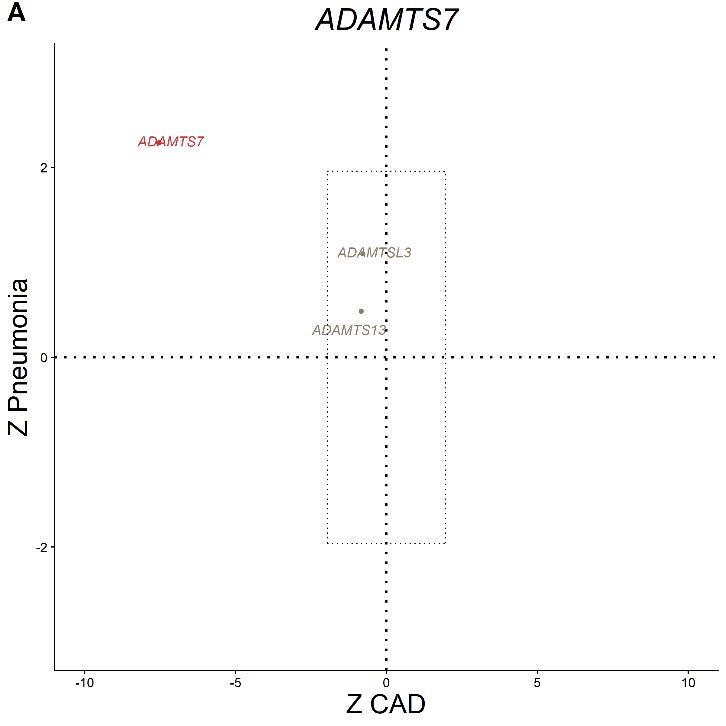 | 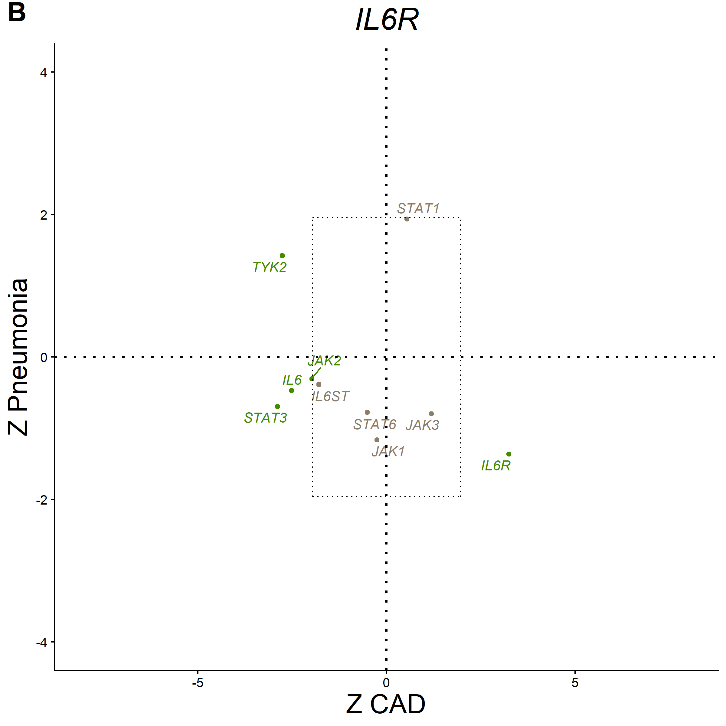 |
| --- | --- |
| 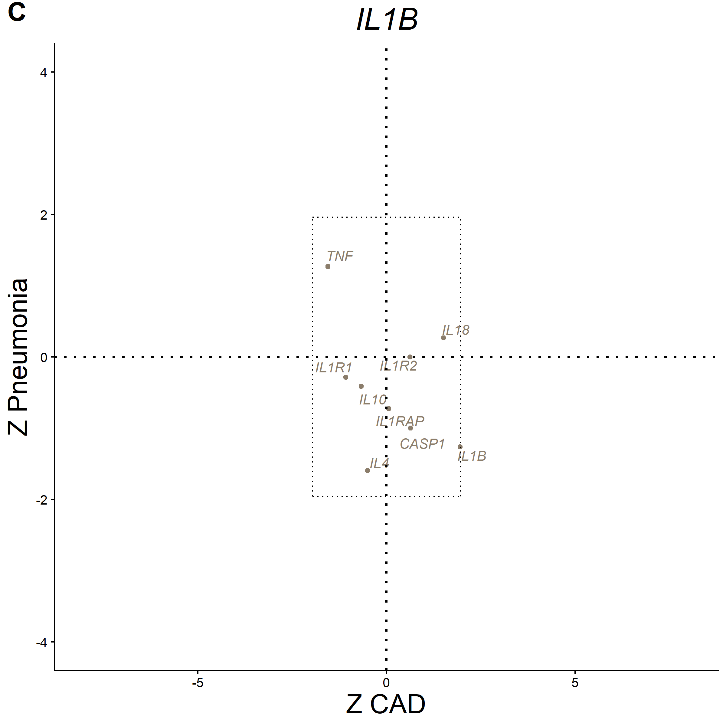 | 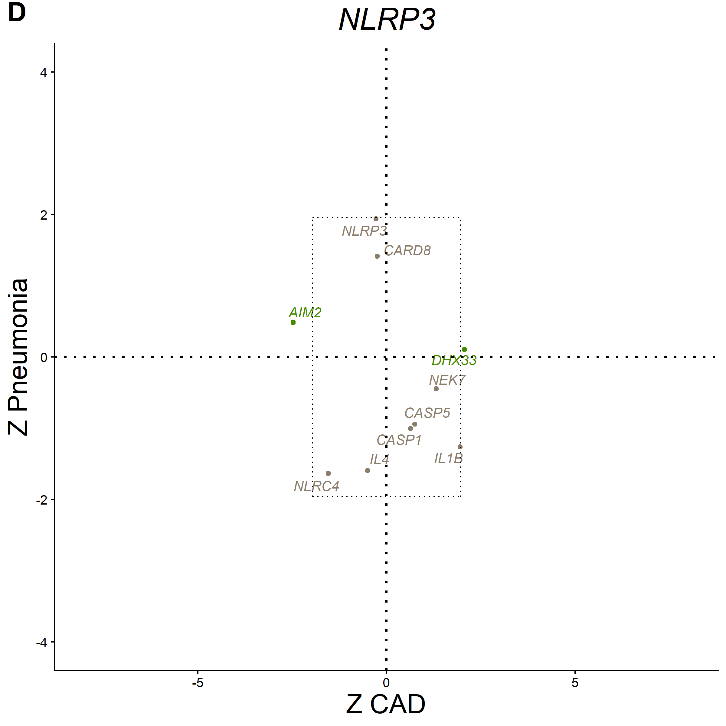 |

###### Supplemental Figure 5. Causal effects of interacting genes with (A) *ADAMTS7*, (B) *IL6R*, (C) *IL1B*, and (D) *NLRP3* on CAD and pneumonia using summary data–based Mendelian randomization methods. eQTL summary statistics for *ADAMTS7* related biomarkers were from artery – tibial tissue of the GTEx v7. Cis-eQTL summary statistics for *IL6R, IL1B, and NLRP3* related biomarkers were from whole blood of the eQTLGen consortium. Mendelian randomization with summary data–based Mendelian randomization (SMR) methods were used for this analysis. Red dots indicate significant causal relations with both CAD and pneumonia. Green dots indicate significant causal relations with either CAD or pneumonia. Grey dots indicate significant causal relations with neither CAD nor pneumonia. Black dashed lines indicate Z scores of 1.96, corresponding to p=0.05. CAD: coronary artery disease, eQTL: expression quantitative trait loci.

### **Supplemental Table 1. Details of 115 SNPs with genetic pleiotropy.**

| **CHR** | **BP** | **Nearest gene** | **RSID** | **ALT** | **REF** | **EAF** | **Pneumonia** | | **CAD** | |
| --- | --- | --- | --- | --- | --- | --- | --- | --- | --- | --- |
|  |  |  |  |  |  |  | **Beta (SE)** | **P** | **Beta (SE)** | **P** |
| 1 | 154395125 | *IL6R* | rs11265611 | A | G | 0.57 | 0.016 (0.01) | 9.87E-02 | -0.024 (0.01) | 1.51E-02 |
| 1 | 154395212 | *IL6R* | rs4133213 | A | C | 0.45 | 0.018 (0.01) | 6.50E-02 | -0.027 (0.01) | 5.79E-03 |
| 1 | 154395839 | *IL6R* | rs6684439 | T | C | 0.41 | 0.023 (0.01) | 1.77E-02 | -0.031 (0.01) | 1.90E-03 |
| 1 | 154395946 | *IL6R* | rs6689306 | A | G | 0.45 | -0.019 (0.01) | 4.52E-02 | 0.027 (0.01) | 7.44E-03 |
| 1 | 154397416 | *IL6R* | rs12118721 | T | C | 0.45 | -0.021 (0.01) | 3.18E-02 | 0.025 (0.01) | 1.12E-02 |
| 1 | 154397610 | *IL6R* | rs12117832 | A | G | 0.45 | -0.021 (0.01) | 3.17E-02 | 0.025 (0.01) | 1.09E-02 |
| 1 | 154400015 | *IL6R* | rs4845618 | T | G | 0.55 | 0.02 (0.01) | 3.36E-02 | -0.026 (0.01) | 9.56E-03 |
| 1 | 154400320 | *IL6R* | rs6687726 | A | G | 0.45 | -0.021 (0.01) | 3.02E-02 | 0.026 (0.01) | 9.88E-03 |
| 1 | 154405024 | *IL6R* | rs56383622 | A | G | 0.58 | -0.019 (0.01) | 4.26E-02 | 0.034 (0.01) | 6.91E-04 |
| 1 | 154406656 | *IL6R* | rs4845620 | A | G | 0.58 | -0.019 (0.01) | 4.46E-02 | 0.034 (0.01) | 6.29E-04 |
| 1 | 154407419 | *IL6R* | rs7518199 | A | C | 0.58 | -0.019 (0.01) | 4.67E-02 | 0.034 (0.01) | 6.06E-04 |
| 1 | 154407713 | *IL6R* | rs7521458 | T | C | 0.58 | -0.019 (0.01) | 4.71E-02 | 0.034 (0.01) | 5.66E-04 |
| 1 | 154409730 | *IL6R* | rs4845621 | A | G | 0.42 | 0.019 (0.01) | 4.37E-02 | -0.034 (0.01) | 5.53E-04 |
| 1 | 154411419 | *IL6R* | rs4845622 | A | C | 0.58 | -0.02 (0.01) | 4.02E-02 | 0.034 (0.01) | 5.45E-04 |
| 1 | 154414037 | *IL6R* | rs4393147 | T | C | 0.42 | 0.02 (0.01) | 4.14E-02 | -0.034 (0.01) | 5.76E-04 |
| 1 | 154414086 | *IL6R* | rs4453032 | A | G | 0.58 | -0.02 (0.01) | 3.98E-02 | 0.034 (0.01) | 5.76E-04 |
| 1 | 154414296 | *IL6R* | rs6664201 | T | C | 0.42 | 0.02 (0.01) | 3.97E-02 | -0.034 (0.01) | 5.76E-04 |
| 1 | 154415396 | *IL6R* | rs4845372 | A | C | 0.43 | 0.018 (0.01) | 5.52E-02 | -0.036 (0.01) | 2.76E-04 |
| 1 | 154415777 | *IL6R* | rs4845623 | A | G | 0.57 | -0.02 (0.011) | 6.46E-02 | 0.042 (0.011) | 1.34E-04 |
| 1 | 154416935 | *IL6R* | rs12753254 | A | G | 0.42 | 0.02 (0.01) | 3.37E-02 | -0.035 (0.01) | 4.42E-04 |
| 1 | 154416969 | *IL6R* | rs12730036 | T | C | 0.42 | 0.02 (0.01) | 3.48E-02 | -0.035 (0.01) | 4.24E-04 |
| 1 | 154417829 | *IL6R* | rs4845373 | T | C | 0.42 | 0.019 (0.01) | 4.66E-02 | -0.034 (0.01) | 5.94E-04 |
| 1 | 154418749 | *IL6R* | rs4576655 | T | C | 0.42 | 0.019 (0.01) | 5.12E-02 | -0.035 (0.01) | 4.44E-04 |
| 1 | 154418879 | *IL6R* | rs4537545 | T | C | 0.42 | 0.019 (0.01) | 4.87E-02 | -0.035 (0.01) | 4.81E-04 |
| 1 | 154420087 | *IL6R* | rs61812598 | A | G | 0.41 | 0.02 (0.01) | 3.66E-02 | -0.033 (0.01) | 1.08E-03 |
| 1 | 154420402 | *IL6R* | rs7512646 | C | G | 0.42 | 0.019 (0.01) | 4.94E-02 | -0.034 (0.01) | 5.90E-04 |
| 1 | 154420778 | *IL6R* | rs7529229 | T | C | 0.58 | -0.019 (0.01) | 5.10E-02 | 0.034 (0.01) | 6.20E-04 |
| 1 | 154425456 | *IL6R* | rs12126142 | A | G | 0.41 | 0.02 (0.01) | 3.97E-02 | -0.033 (0.01) | 7.77E-04 |
| 1 | 154426264 | *IL6R* | rs4129267 | T | C | 0.41 | 0.02 (0.01) | 4.22E-02 | -0.033 (0.01) | 7.83E-04 |
| 1 | 154426970 | *IL6R* | rs2228145 | A | C | 0.59 | -0.02 (0.011) | 5.55E-02 | 0.042 (0.011) | 1.34E-04 |
| 1 | 154428283 | *IL6R* | rs12133641 | A | G | 0.59 | -0.02 (0.01) | 4.00E-02 | 0.034 (0.01) | 6.30E-04 |
| 12 | 111865049 | *SH2B3* | rs7310615 | C | G | 0.49 | -0.003 (0.01) | 7.35E-01 | 0.076 (0.01) | 1.97E-14 |
| 12 | 112007756 | *ATXN2* | rs653178 | T | C | 0.51 | 0.004 (0.01) | 6.37E-01 | -0.076 (0.01) | 1.46E-14 |
| 15 | 79031381 | *CHRNB4* | rs12907372 | NA | NA | NA | NA | NA | NA | NA |
| 15 | 79033520 | *CHRNB4* | rs56354501 | A | G | 0.60 | -0.041 (0.01) | 3.04E-05 | 0.071 (0.01) | 1.69E-12 |
| 15 | 79034276 | *CHRNB4* | rs8031513 | T | C | 0.58 | -0.048 (0.011) | 8.22E-06 | 0.068 (0.011) | 6.34E-10 |
| 15 | 79043402 | *ADAMTS7* | rs4887090 | A | G | 0.42 | 0.048 (0.011) | 5.89E-06 | NA | NA |
| 15 | 79049766 | *ADAMTS7* | rs55834964 | A | C | 0.42 | 0.047 (0.011) | 9.95E-06 | -0.071 (0.011) | 1.09E-10 |
| 15 | 79051759 | *ADAMTS7* | rs12286 | A | G | 0.42 | 0.048 (0.011) | 7.51E-06 | -0.072 (0.011) | 5.93E-11 |
| 15 | 79052277 | *ADAMTS7* | rs7182642 | T | C | 0.40 | 0.039 (0.01) | 4.93E-05 | -0.079 (0.01) | 4.65E-15 |
| 15 | 79052580 | *ADAMTS7* | rs12906653 | A | G | 0.42 | 0.047 (0.011) | 8.96E-06 | -0.074 (0.011) | 1.73E-11 |
| 15 | 79053207 | *ADAMTS7* | rs4887095 | A | C | 0.42 | 0.049 (0.011) | 3.55E-06 | NA | NA |
| 15 | 79053284 | *ADAMTS7* | rs4887096 | T | C | 0.57 | -0.048 (0.011) | 5.61E-06 | 0.074 (0.011) | 1.73E-11 |
| 15 | 79053814 | *ADAMTS7* | rs1809419 | A | G | 0.42 | 0.048 (0.011) | 5.77E-06 | -0.074 (0.011) | 1.73E-11 |
| 15 | 79054011 | *ADAMTS7* | rs12916326 | A | G | 0.57 | -0.048 (0.011) | 6.25E-06 | 0.074 (0.011) | 1.73E-11 |
| 15 | 79054117 | *ADAMTS7* | rs12898346 | T | C | 0.58 | -0.05 (0.011) | 3.50E-06 | NA | NA |
| 15 | 79054129 | *ADAMTS7* | rs12916648 | T | C | 0.42 | 0.049 (0.011) | 5.00E-06 | -0.074 (0.011) | 1.73E-11 |
| 15 | 79055163 | *ADAMTS7* | rs7168391 | T | G | 0.41 | 0.039 (0.01) | 5.34E-05 | -0.106 (0.024) | 1.37E-05 |
| 15 | 79056769 | *ADAMTS7* | rs1809420 | T | C | 0.57 | -0.047 (0.011) | 1.03E-05 | 0.075 (0.011) | 9.22E-12 |
| 15 | 79056815 | *ADAMTS7* | rs11633351 | T | C | 0.43 | 0.047 (0.011) | 1.08E-05 | -0.075 (0.011) | 9.22E-12 |
| 15 | 79057093 | *ADAMTS7* | rs6495267 | A | G | 0.43 | 0.047 (0.011) | 1.03E-05 | -0.075 (0.011) | 9.22E-12 |
| 15 | 79057949 | *ADAMTS7* | rs4887097 | T | C | 0.57 | -0.047 (0.011) | 9.07E-06 | NA | NA |
| 15 | 79057950 | *ADAMTS7* | rs4887098 | C | G | 0.43 | 0.047 (0.011) | 9.06E-06 | NA | NA |
| 15 | 79058951 | *ADAMTS7* | rs28699256 | T | C | 0.54 | -0.047 (0.011) | 8.39E-06 | 0.068 (0.011) | 6.34E-10 |
| 15 | 79059523 | *ADAMTS7* | rs3894351 | A | C | 0.41 | 0.042 (0.01) | 1.55E-05 | -0.105 (0.024) | 1.60E-05 |
| 15 | 79059526 | *ADAMTS7* | rs3894352 | T | G | 0.59 | -0.042 (0.01) | 1.47E-05 | 0.105 (0.024) | 1.65E-05 |
| 15 | 79059547 | *ADAMTS7* | rs4887099 | C | G | 0.41 | 0.042 (0.01) | 1.54E-05 | -0.104 (0.024) | 1.86E-05 |
| 15 | 79059670 | *ADAMTS7* | rs1809423 | T | C | 0.59 | -0.042 (0.01) | 1.59E-05 | 0.105 (0.024) | 1.51E-05 |
| 15 | 79059691 | *ADAMTS7* | rs2904223 | A | G | 0.41 | 0.039 (0.01) | 5.03E-05 | -0.08 (0.01) | 1.36E-15 |
| 15 | 79061002 | *ADAMTS7* | rs8043119 | A | G | 0.43 | 0.047 (0.011) | 8.39E-06 | -0.075 (0.011) | 9.22E-12 |
| 15 | 79062102 | *ADAMTS7* | rs1807007 | T | G | 0.57 | -0.046 (0.011) | 1.40E-05 | 0.075 (0.011) | 9.22E-12 |
| 15 | 79062340 | *ADAMTS7* | rs1807006 | C | G | 0.57 | -0.046 (0.011) | 1.39E-05 | 0.075 (0.011) | 9.22E-12 |
| 15 | 79063474 | *ADAMTS7* | rs1809409 | A | T | 0.57 | -0.046 (0.011) | 1.39E-05 | 0.075 (0.011) | 9.22E-12 |
| 15 | 79064080 | *ADAMTS7* | rs11635931 | A | G | 0.43 | 0.046 (0.011) | 1.39E-05 | -0.075 (0.011) | 9.22E-12 |
| 15 | 79064143 | *ADAMTS7* | rs11635870 | C | G | 0.57 | -0.046 (0.011) | 1.39E-05 | 0.075 (0.011) | 9.22E-12 |
| 15 | 79064667 | *ADAMTS7* | rs7174367 | A | G | 0.57 | -0.046 (0.011) | 1.92E-05 | 0.075 (0.011) | 9.22E-12 |
| 15 | 79067385 | *ADAMTS7* | rs35934157 | A | G | 0.41 | 0.044 (0.011) | 4.59E-05 | -0.073 (0.011) | 3.22E-11 |
| 15 | 79067922 | *ADAMTS7* | rs7171578 | T | C | 0.41 | 0.045 (0.011) | 2.81E-05 | -0.074 (0.011) | 1.73E-11 |
| 15 | 79067951 | *ADAMTS7* | rs7171916 | C | G | 0.54 | -0.037 (0.01) | 1.52E-04 | 0.081 (0.01) | 6.36E-16 |
| 15 | 79068328 | *ADAMTS7* | rs12906691 | A | G | 0.41 | 0.044 (0.011) | 4.25E-05 | -0.074 (0.011) | 1.73E-11 |
| 15 | 79069121 | *ADAMTS7* | rs11854507 | A | G | 0.59 | -0.043 (0.011) | 5.89E-05 | 0.074 (0.011) | 1.73E-11 |
| 15 | 79069734 | *ADAMTS7* | rs7161774 | T | G | 0.55 | -0.039 (0.01) | 6.30E-05 | 0.081 (0.01) | 8.55E-16 |
| 15 | 79070438 | *ADAMTS7* | rs12907764 | A | G | 0.59 | -0.026 (0.01) | 8.25E-03 | 0.078 (0.01) | 8.94E-15 |
| 15 | 79071095 | *ADAMTS7* | rs12913260 | A | G | 0.42 | 0.029 (0.01) | 2.31E-03 | -0.072 (0.01) | 5.65E-13 |
| 15 | 79071406 | *ADAMTS7* | rs2004038 | A | G | 0.42 | 0.029 (0.01) | 2.87E-03 | -0.071 (0.01) | 8.68E-13 |
| 15 | 79072100 | *ADAMTS7* | rs112321636 | T | G | 0.42 | 0.029 (0.01) | 2.34E-03 | -0.071 (0.01) | 1.16E-12 |
| 15 | 79074000 | *ADAMTS7* | rs35474770 | A | G | 0.58 | -0.03 (0.01) | 2.03E-03 | 0.071 (0.01) | 7.43E-13 |
| 15 | 79074253 | *ADAMTS7* | rs36061084 | A | G | 0.42 | 0.029 (0.01) | 2.10E-03 | -0.071 (0.01) | 7.27E-13 |
| 15 | 79074294 | *ADAMTS7* | rs4887109 | T | C | 0.42 | 0.03 (0.01) | 1.90E-03 | -0.069 (0.01) | 3.13E-12 |
| 15 | 79074506 | *ADAMTS7* | rs4886590 | A | G | 0.42 | 0.03 (0.01) | 1.57E-03 | -0.07 (0.01) | 1.72E-12 |
| 15 | 79074518 | *ADAMTS7* | rs4886591 | A | G | 0.45 | 0.034 (0.01) | 3.02E-04 | -0.071 (0.01) | 9.16E-13 |
| 15 | 79075335 | *ADAMTS7* | rs10163145 | A | G | 0.44 | 0.03 (0.01) | 1.76E-03 | -0.065 (0.023) | 4.56E-03 |
| 15 | 79075746 | *ADAMTS7* | rs12050525 | T | C | 0.56 | -0.029 (0.01) | 2.55E-03 | 0.069 (0.01) | 2.83E-12 |
| 15 | 79077114 | *ADAMTS7* | rs1825087 | A | G | 0.56 | -0.029 (0.01) | 2.67E-03 | 0.069 (0.01) | 2.83E-12 |
| 15 | 79079074 | *ADAMTS7* | rs11072806 | A | G | 0.44 | 0.028 (0.01) | 3.35E-03 | -0.068 (0.01) | 5.39E-12 |
| 15 | 79079512 | *ADAMTS7* | rs12899147 | A | G | 0.56 | -0.029 (0.01) | 2.71E-03 | 0.069 (0.01) | 4.79E-12 |
| 15 | 79080234 | *ADAMTS7* | rs1994016 | T | C | 0.42 | 0.023 (0.01) | 1.49E-02 | -0.065 (0.01) | 7.84E-11 |
| 15 | 79082547 | *ADAMTS7* | rs4886592 | T | C | 0.56 | -0.03 (0.01) | 1.84E-03 | 0.068 (0.01) | 6.43E-12 |
| 15 | 79083376 | *ADAMTS7* | rs2277546 | A | C | 0.44 | 0.03 (0.01) | 1.72E-03 | -0.068 (0.01) | 6.39E-12 |
| 15 | 79083591 | *ADAMTS7* | rs2277545 | T | C | 0.56 | -0.03 (0.01) | 1.74E-03 | 0.068 (0.01) | 6.50E-12 |
| 15 | 79083814 | *ADAMTS7* | rs11639044 | T | C | 0.44 | 0.03 (0.01) | 1.53E-03 | -0.068 (0.01) | 1.03E-11 |
| 15 | 79084933 | *ADAMTS7* | rs12903203 | T | C | 0.56 | -0.031 (0.01) | 1.38E-03 | 0.068 (0.01) | 7.38E-12 |
| 15 | 79085915 | *ADAMTS7* | rs11631955 | A | G | 0.56 | -0.031 (0.01) | 9.83E-04 | 0.067 (0.01) | 1.69E-11 |
| 15 | 79086057 | *ADAMTS7* | rs11632102 | A | G | 0.44 | 0.032 (0.01) | 7.62E-04 | -0.066 (0.01) | 2.11E-11 |
| 15 | 79089111 | *ADAMTS7* | rs3825807 | A | G | 0.55 | -0.027 (0.01) | 4.77E-03 | 0.069 (0.01) | 3.20E-12 |
| 15 | 79089734 | *ADAMTS7* | rs7177699 | T | C | 0.55 | -0.027 (0.01) | 4.46E-03 | 0.069 (0.01) | 3.39E-12 |
| 15 | 79090606 | *ADAMTS7* | rs28610385 | A | C | 0.45 | 0.027 (0.01) | 4.22E-03 | -0.069 (0.01) | 3.04E-12 |
| 15 | 79092750 | *ADAMTS7* | rs7173267 | C | G | 0.45 | 0.027 (0.01) | 4.47E-03 | -0.069 (0.01) | 2.91E-12 |
| 15 | 79093201 | *ADAMTS7* | rs11634450 | A | G | 0.55 | -0.027 (0.01) | 4.34E-03 | 0.069 (0.01) | 3.25E-12 |
| 15 | 79094325 | *ADAMTS7* | rs11856536 | A | G | 0.55 | -0.027 (0.01) | 4.20E-03 | 0.069 (0.01) | 3.23E-12 |
| 15 | 79095287 | *ADAMTS7* | rs4887113 | T | C | 0.45 | 0.028 (0.01) | 3.56E-03 | -0.067 (0.01) | 1.17E-11 |
| 15 | 79099145 | *ADAMTS7* | rs12592721 | C | G | 0.45 | 0.027 (0.01) | 4.23E-03 | -0.069 (0.01) | 3.84E-12 |
| 15 | 79105350 | *MORF4L1* | rs11634042 | T | C | 0.44 | 0.023 (0.01) | 1.61E-02 | -0.069 (0.01) | 2.57E-12 |
| 15 | 79114453 | *MORF4L1* | rs28455815 | T | C | 0.55 | -0.016 (0.01) | 9.03E-02 | 0.073 (0.01) | 2.42E-13 |
| 15 | 79121776 | *MORF4L1* | rs7179953 | T | C | 0.55 | -0.016 (0.01) | 9.06E-02 | 0.073 (0.01) | 2.31E-13 |
| 15 | 79123054 | *MORF4L1* | rs7164479 | T | C | 0.55 | -0.017 (0.01) | 7.85E-02 | 0.072 (0.01) | 4.32E-13 |
| 15 | 79123338 | *MORF4L1* | rs7165042 | C | G | 0.45 | 0.016 (0.01) | 8.85E-02 | -0.073 (0.01) | 2.31E-13 |
| 15 | 79123396 | *MORF4L1* | rs7165081 | C | G | 0.45 | 0.016 (0.01) | 8.86E-02 | -0.073 (0.01) | 2.31E-13 |
| 15 | 79123505 | *MORF4L1* | rs7165733 | A | G | 0.55 | -0.016 (0.01) | 9.04E-02 | 0.073 (0.01) | 2.31E-13 |
| 15 | 79123509 | *MORF4L1* | rs7166764 | T | G | 0.45 | 0.016 (0.01) | 9.04E-02 | -0.073 (0.01) | 2.31E-13 |
| 15 | 79123631 | *MORF4L1* | rs7182529 | T | C | 0.45 | 0.016 (0.01) | 9.10E-02 | -0.073 (0.01) | 2.31E-13 |
| 15 | 79123753 | *MORF4L1* | rs7182716 | T | C | 0.45 | 0.016 (0.01) | 9.36E-02 | -0.073 (0.01) | 2.31E-13 |
| 15 | 79123779 | *MORF4L1* | rs7181432 | T | C | 0.55 | -0.016 (0.01) | 9.34E-02 | 0.073 (0.01) | 2.31E-13 |
| 15 | 79123946 | *MORF4L1* | rs7182103 | T | G | 0.55 | -0.016 (0.01) | 9.77E-02 | 0.073 (0.01) | 2.31E-13 |
| 15 | 79124475 | *MORF4L1* | rs4468572 | T | C | 0.45 | 0.016 (0.01) | 9.92E-02 | -0.073 (0.01) | 2.19E-13 |

Genetic pleiotropy defined as PLACO p values below genome-wide significance (5E-8) with input GWAS of CAD from CARDIoGRAMplusC4D and that of pneumonia from FinnGen R4. Effect size was evaluated in GWAS from meta-analysis of UK Biobank and Mass General Brigham Biobank. CAD: coronary artery disease, EAF: effect allele frequency, GWAS: genome-wide association study.

### **Supplemental Table 2. Causal effects of the expression of genes interacting with *ADAMTS7* on CAD and pneumonia.**

| **Exposure** | **Outcome** | **Methods** | **No of SNPs** | **Beta** | **SE** | **P** |
| --- | --- | --- | --- | --- | --- | --- |
| *ADAMTS13* | CAD | Single SNP | 1 | -0.016 | 0.019 | 4.03E-01 |
|  | Pneumonia | Single SNP | 1 | 0.013 | 0.028 | 6.28E-01 |
| *ADAMTS7* | CAD | Inverse variance weighted (fixed effects) | 2 | -0.161 | 0.015 | 9.12E-28 |
|  |  | MR RAPS | 2 | -0.164 | 0.019 | 1.65E-18 |
|  | Pneumonia | Inverse variance weighted (fixed effects) | 2 | 0.066 | 0.021 | 1.34E-03 |
|  |  | MR RAPS | 2 | 0.066 | 0.022 | 2.22E-03 |
| *ADAMTSL1* | CAD | Single SNP | 1 | -0.009 | 0.029 | 7.66E-01 |
|  | Pneumonia | Single SNP | 1 | 0.000 | 0.040 | 9.91E-01 |
| *ADAMTSL3* | CAD | Single SNP | 1 | -0.017 | 0.022 | 4.34E-01 |
|  | Pneumonia | Single SNP | 1 | 0.033 | 0.030 | 2.69E-01 |

###### eQTL data derived from tibial artery tissue for *ADAMTS7* related genes were from the GTEx v8. Mendelian randomization with either Wald ratio method (single instrument) or inverse-variance weighted and MR RAPS methods (two instruments) were used for this analysis. CAD: coronary artery disease, eQTL: expression quantitative trait loci.

### **Supplemental Table 3. Causal effects of the expression of genes interacting with *IL6R, IL1B,* or *NLRP3* on CAD and pneumonia.**

| **Exposure** | **Outcome** | **Methods** | **No of SNPs** | **Beta** | **SE** | **P** |
| --- | --- | --- | --- | --- | --- | --- |
| ***IL6R*** | | | | | | |
| *IL6* | CAD | Inverse variance weighted (fixed effects) | 3 | -0.209 | 0.090 | 2.07E-02 |
|  |  | MR Egger | 3 | -0.212 | 0.822 | 8.39E-01 |
|  |  | MR RAPS | 3 | -0.214 | 0.094 | 2.27E-02 |
|  |  | Weighted median | 3 | -0.205 | 0.108 | 5.78E-02 |
|  |  | Weighted mode | 3 | -0.096 | 0.120 | 5.04E-01 |
|  | Pneumonia | Inverse variance weighted (fixed effects) | 3 | -0.202 | 0.122 | 9.85E-02 |
|  |  | MR Egger | 3 | 0.448 | 0.579 | 5.81E-01 |
|  |  | MR RAPS | 3 | -0.203 | 0.125 | 1.06E-01 |
|  |  | Weighted median | 3 | -0.132 | 0.136 | 3.33E-01 |
|  |  | Weighted mode | 3 | -0.124 | 0.161 | 5.23E-01 |
| *IL6R* | CAD | Inverse variance weighted (fixed effects) | 5 | 0.280 | 0.079 | 3.76E-04 |
|  |  | MR Egger | 5 | 0.262 | 0.463 | 6.12E-01 |
|  |  | MR RAPS | 5 | 0.287 | 0.081 | 3.99E-04 |
|  |  | Weighted median | 5 | 0.219 | 0.096 | 2.29E-02 |
|  |  | Weighted mode | 5 | 0.334 | 0.110 | 3.81E-02 |
|  | Pneumonia | Inverse variance weighted (fixed effects) | 5 | -0.237 | 0.108 | 2.87E-02 |
|  |  | MR Egger | 5 | 0.300 | 0.549 | 6.22E-01 |
|  |  | MR RAPS | 5 | -0.242 | 0.110 | 2.83E-02 |
|  |  | Weighted median | 5 | -0.208 | 0.123 | 9.01E-02 |
|  |  | Weighted mode | 5 | -0.187 | 0.131 | 2.28E-01 |
| *IL6ST* | CAD | Inverse variance weighted (fixed effects) | 4 | -0.096 | 0.059 | 1.06E-01 |
|  |  | MR Egger | 4 | -0.318 | 0.207 | 2.64E-01 |
|  |  | MR RAPS | 4 | -0.097 | 0.061 | 1.11E-01 |
|  |  | Weighted median | 4 | -0.118 | 0.070 | 9.10E-02 |
|  |  | Weighted mode | 4 | -0.135 | 0.074 | 1.63E-01 |
|  | Pneumonia | Inverse variance weighted (fixed effects) | 4 | -0.099 | 0.082 | 2.24E-01 |
|  |  | MR Egger | 4 | -0.046 | 0.345 | 9.07E-01 |
|  |  | MR RAPS | 4 | -0.100 | 0.083 | 2.28E-01 |
|  |  | Weighted median | 4 | -0.057 | 0.098 | 5.63E-01 |
|  |  | Weighted mode | 4 | -0.046 | 0.115 | 7.14E-01 |
| *STAT1* | CAD | Inverse variance weighted (fixed effects) | 11 | 0.224 | 0.035 | 8.66E-11 |
|  |  | MR Egger | 11 | 0.603 | 0.205 | 1.65E-02 |
|  |  | MR RAPS | 11 | 0.245 | 0.034 | 1.19E-12 |
|  |  | Weighted median | 11 | 0.044 | 0.070 | 5.32E-01 |
|  |  | Weighted mode | 11 | -0.011 | 0.076 | 8.83E-01 |
|  | Pneumonia | Inverse variance weighted (fixed effects) | 11 | -0.041 | 0.052 | 4.32E-01 |
|  |  | MR Egger | 11 | -0.056 | 0.201 | 7.85E-01 |
|  |  | MR RAPS | 11 | -0.042 | 0.052 | 4.19E-01 |
|  |  | Weighted median | 11 | 0.027 | 0.073 | 7.17E-01 |
|  |  | Weighted mode | 11 | 0.080 | 0.089 | 3.91E-01 |
| *STAT3* | CAD | Inverse variance weighted (fixed effects) | 7 | -0.250 | 0.048 | 1.90E-07 |
|  |  | MR Egger | 7 | 0.015 | 0.204 | 9.43E-01 |
|  |  | MR RAPS | 7 | -0.255 | 0.049 | 2.61E-07 |
|  |  | Weighted median | 7 | -0.216 | 0.059 | 2.34E-04 |
|  |  | Weighted mode | 7 | -0.208 | 0.058 | 1.11E-02 |
|  | Pneumonia | Inverse variance weighted (fixed effects) | 8 | -0.017 | 0.065 | 7.96E-01 |
|  |  | MR Egger | 8 | -0.128 | 0.173 | 4.87E-01 |
|  |  | MR RAPS | 8 | -0.017 | 0.065 | 7.97E-01 |
|  |  | Weighted median | 8 | -0.062 | 0.078 | 4.30E-01 |
|  |  | Weighted mode | 8 | -0.056 | 0.080 | 5.03E-01 |
| *STAT6* | CAD | Inverse variance weighted (fixed effects) | 37 | -0.017 | 0.020 | 3.80E-01 |
|  |  | MR Egger | 37 | -0.031 | 0.051 | 5.48E-01 |
|  |  | MR RAPS | 37 | -0.017 | 0.020 | 3.76E-01 |
|  |  | Weighted median | 37 | -0.018 | 0.028 | 5.11E-01 |
|  |  | Weighted mode | 37 | -0.021 | 0.027 | 4.45E-01 |
|  | Pneumonia | Inverse variance weighted (fixed effects) | 42 | 0.026 | 0.026 | 3.10E-01 |
|  |  | MR Egger | 42 | -0.006 | 0.051 | 9.09E-01 |
|  |  | MR RAPS | 42 | 0.026 | 0.026 | 3.10E-01 |
|  |  | Weighted median | 42 | 0.039 | 0.037 | 2.89E-01 |
|  |  | Weighted mode | 42 | 0.023 | 0.032 | 4.73E-01 |
| *TYK2* | CAD | Inverse variance weighted (fixed effects) | 18 | -0.093 | 0.031 | 3.03E-03 |
|  |  | MR Egger | 18 | -0.077 | 0.090 | 4.02E-01 |
|  |  | MR RAPS | 18 | -0.096 | 0.031 | 2.24E-03 |
|  |  | Weighted median | 18 | -0.089 | 0.039 | 2.31E-02 |
|  |  | Weighted mode | 18 | -0.093 | 0.039 | 3.04E-02 |
|  | Pneumonia | Inverse variance weighted (fixed effects) | 18 | 0.069 | 0.044 | 1.17E-01 |
|  |  | MR Egger | 18 | 0.018 | 0.098 | 8.53E-01 |
|  |  | MR RAPS | 18 | 0.070 | 0.044 | 1.17E-01 |
|  |  | Weighted median | 18 | 0.079 | 0.054 | 1.45E-01 |
|  |  | Weighted mode | 18 | 0.056 | 0.053 | 3.05E-01 |
| ***IL1B*** | | | | | | |
| *CASP1* | CAD | Inverse variance weighted (fixed effects) | 9 | -0.031 | 0.045 | 4.91E-01 |
|  |  | MR Egger | 9 | 0.101 | 0.116 | 4.12E-01 |
|  |  | MR RAPS | 9 | -0.032 | 0.045 | 4.86E-01 |
|  |  | Weighted median | 9 | -0.007 | 0.061 | 9.12E-01 |
|  |  | Weighted mode | 9 | 0.067 | 0.058 | 2.83E-01 |
|  | Pneumonia | Inverse variance weighted (fixed effects) | 9 | -0.073 | 0.062 | 2.41E-01 |
|  |  | MR Egger | 9 | 0.016 | 0.114 | 8.92E-01 |
|  |  | MR RAPS | 9 | -0.073 | 0.063 | 2.44E-01 |
|  |  | Weighted median | 9 | -0.082 | 0.072 | 2.54E-01 |
|  |  | Weighted mode | 9 | -0.066 | 0.077 | 4.19E-01 |
| *IL10* | CAD | Inverse variance weighted (fixed effects) | 5 | -0.102 | 0.047 | 3.20E-02 |
|  |  | MR Egger | 5 | -0.062 | 0.175 | 7.46E-01 |
|  |  | MR RAPS | 5 | -0.104 | 0.048 | 2.97E-02 |
|  |  | Weighted median | 5 | -0.134 | 0.057 | 1.87E-02 |
|  |  | Weighted mode | 5 | -0.113 | 0.057 | 1.18E-01 |
|  | Pneumonia | Inverse variance weighted (fixed effects) | 5 | -0.115 | 0.070 | 9.78E-02 |
|  |  | MR Egger | 5 | 0.062 | 0.141 | 6.91E-01 |
|  |  | MR RAPS | 5 | -0.117 | 0.070 | 9.67E-02 |
|  |  | Weighted median | 5 | -0.028 | 0.078 | 7.22E-01 |
|  |  | Weighted mode | 5 | -0.026 | 0.088 | 7.78E-01 |
| *IL18* | CAD | Inverse variance weighted (fixed effects) | 9 | 0.011 | 0.030 | 7.05E-01 |
|  |  | MR Egger | 9 | -0.013 | 0.056 | 8.28E-01 |
|  |  | MR RAPS | 9 | 0.011 | 0.030 | 7.08E-01 |
|  |  | Weighted median | 9 | 0.009 | 0.033 | 7.73E-01 |
|  |  | Weighted mode | 9 | 0.006 | 0.034 | 8.55E-01 |
|  | Pneumonia | Inverse variance weighted (fixed effects) | 9 | 0.011 | 0.041 | 7.96E-01 |
|  |  | MR Egger | 9 | -0.039 | 0.076 | 6.28E-01 |
|  |  | MR RAPS | 9 | 0.011 | 0.041 | 7.96E-01 |
|  |  | Weighted median | 9 | 0.010 | 0.046 | 8.19E-01 |
|  |  | Weighted mode | 9 | -0.003 | 0.043 | 9.54E-01 |
| *IL1B* | CAD | Inverse variance weighted (fixed effects) | 10 | 0.058 | 0.050 | 2.42E-01 |
|  |  | MR Egger | 10 | 0.831 | 0.281 | 1.82E-02 |
|  |  | MR RAPS | 10 | 0.061 | 0.050 | 2.20E-01 |
|  |  | Weighted median | 10 | 0.168 | 0.075 | 2.45E-02 |
|  |  | Weighted mode | 10 | 0.186 | 0.079 | 4.24E-02 |
|  | Pneumonia | Inverse variance weighted (fixed effects) | 10 | 0.084 | 0.069 | 2.22E-01 |
|  |  | MR Egger | 10 | 0.328 | 0.342 | 3.66E-01 |
|  |  | MR RAPS | 10 | 0.086 | 0.070 | 2.20E-01 |
|  |  | Weighted median | 10 | 0.155 | 0.098 | 1.11E-01 |
|  |  | Weighted mode | 10 | 0.181 | 0.137 | 2.18E-01 |
| *IL1R1* | CAD | Inverse variance weighted (fixed effects) | 5 | -0.010 | 0.051 | 8.44E-01 |
|  |  | MR Egger | 5 | -0.017 | 0.353 | 9.64E-01 |
|  |  | MR RAPS | 5 | -0.010 | 0.051 | 8.41E-01 |
|  |  | Weighted median | 5 | 0.006 | 0.070 | 9.32E-01 |
|  |  | Weighted mode | 5 | 0.058 | 0.074 | 4.77E-01 |
|  | Pneumonia | Inverse variance weighted (fixed effects) | 5 | 0.015 | 0.068 | 8.24E-01 |
|  |  | MR Egger | 5 | -0.205 | 0.217 | 4.14E-01 |
|  |  | MR RAPS | 5 | 0.015 | 0.068 | 8.25E-01 |
|  |  | Weighted median | 5 | -0.016 | 0.078 | 8.35E-01 |
|  |  | Weighted mode | 5 | -0.021 | 0.089 | 8.27E-01 |
| *IL1R2* | CAD | Inverse variance weighted (fixed effects) | 21 | -0.013 | 0.027 | 6.48E-01 |
|  |  | MR Egger | 21 | -0.009 | 0.050 | 8.66E-01 |
|  |  | MR RAPS | 21 | -0.013 | 0.028 | 6.49E-01 |
|  |  | Weighted median | 21 | 0.002 | 0.035 | 9.53E-01 |
|  |  | Weighted mode | 21 | -0.008 | 0.034 | 8.27E-01 |
|  | Pneumonia | Inverse variance weighted (fixed effects) | 23 | -0.009 | 0.037 | 8.19E-01 |
|  |  | MR Egger | 23 | -0.031 | 0.068 | 6.55E-01 |
|  |  | MR RAPS | 23 | -0.009 | 0.038 | 8.19E-01 |
|  |  | Weighted median | 23 | -0.007 | 0.047 | 8.76E-01 |
|  |  | Weighted mode | 23 | -0.013 | 0.046 | 7.87E-01 |
| *IL1RAP* | CAD | Inverse variance weighted (fixed effects) | 11 | -0.073 | 0.037 | 5.14E-02 |
|  |  | MR Egger | 11 | 0.264 | 0.325 | 4.37E-01 |
|  |  | MR RAPS | 11 | -0.090 | 0.035 | 9.75E-03 |
|  |  | Weighted median | 11 | 0.041 | 0.049 | 4.10E-01 |
|  |  | Weighted mode | 11 | 0.055 | 0.050 | 2.93E-01 |
|  | Pneumonia | Inverse variance weighted (fixed effects) | 11 | 0.000 | 0.050 | 9.95E-01 |
|  |  | MR Egger | 11 | -0.049 | 0.155 | 7.59E-01 |
|  |  | MR RAPS | 11 | 0.000 | 0.050 | 9.95E-01 |
|  |  | Weighted median | 11 | -0.019 | 0.064 | 7.72E-01 |
|  |  | Weighted mode | 11 | -0.015 | 0.082 | 8.61E-01 |
| *IL4* | CAD | Inverse variance weighted (fixed effects) | 4 | 0.008 | 0.071 | 9.06E-01 |
|  |  | MR Egger | 4 | 0.430 | 0.294 | 2.81E-01 |
|  |  | MR RAPS | 4 | 0.009 | 0.071 | 9.03E-01 |
|  |  | Weighted median | 4 | 0.055 | 0.086 | 5.27E-01 |
|  |  | Weighted mode | 4 | 0.121 | 0.085 | 2.51E-01 |
|  | Pneumonia | Inverse variance weighted (fixed effects) | 4 | 0.101 | 0.093 | 2.78E-01 |
|  |  | MR Egger | 4 | 0.433 | 0.246 | 2.20E-01 |
|  |  | MR RAPS | 4 | 0.102 | 0.094 | 2.79E-01 |
|  |  | Weighted median | 4 | 0.175 | 0.099 | 7.65E-02 |
|  |  | Weighted mode | 4 | 0.176 | 0.108 | 2.00E-01 |
| *TNF* | CAD | Inverse variance weighted (fixed effects) | 10 | -0.174 | 0.024 | 3.67E-13 |
|  |  | MR Egger | 10 | -0.015 | 0.071 | 8.34E-01 |
|  |  | MR RAPS | 10 | -0.180 | 0.025 | 3.80E-13 |
|  |  | Weighted median | 10 | -0.164 | 0.028 | 6.63E-09 |
|  |  | Weighted mode | 10 | -0.117 | 0.028 | 2.22E-03 |
|  | Pneumonia | Inverse variance weighted (fixed effects) | 10 | -0.008 | 0.035 | 8.23E-01 |
|  |  | MR Egger | 10 | -0.076 | 0.069 | 2.97E-01 |
|  |  | MR RAPS | 10 | -0.008 | 0.035 | 8.23E-01 |
|  |  | Weighted median | 10 | -0.029 | 0.038 | 4.42E-01 |
|  |  | Weighted mode | 10 | -0.027 | 0.039 | 5.07E-01 |
| ***NLRP3*** | | | | | | |
| *AIM2* | CAD | Inverse variance weighted (fixed effects) | 13 | 0.043 | 0.040 | 2.88E-01 |
|  |  | MR Egger | 13 | -0.400 | 0.268 | 1.63E-01 |
|  |  | MR RAPS | 13 | 0.054 | 0.037 | 1.48E-01 |
|  |  | Weighted median | 13 | -0.134 | 0.058 | 2.08E-02 |
|  |  | Weighted mode | 13 | -0.153 | 0.057 | 2.03E-02 |
|  | Pneumonia | Inverse variance weighted (fixed effects) | 13 | 0.103 | 0.059 | 8.38E-02 |
|  |  | MR Egger | 13 | 0.037 | 0.153 | 8.11E-01 |
|  |  | MR RAPS | 13 | 0.105 | 0.060 | 8.24E-02 |
|  |  | Weighted median | 13 | 0.184 | 0.081 | 2.33E-02 |
|  |  | Weighted mode | 13 | 0.155 | 0.102 | 1.54E-01 |
| *CARD8* | CAD | Inverse variance weighted (fixed effects) | 51 | 0.003 | 0.013 | 8.23E-01 |
|  |  | MR Egger | 51 | -0.003 | 0.021 | 8.87E-01 |
|  |  | MR RAPS | 51 | 0.003 | 0.013 | 8.23E-01 |
|  |  | Weighted median | 51 | 0.011 | 0.020 | 5.89E-01 |
|  |  | Weighted mode | 51 | 0.006 | 0.015 | 7.10E-01 |
|  | Pneumonia | Inverse variance weighted (fixed effects) | 54 | 0.035 | 0.018 | 5.20E-02 |
|  |  | MR Egger | 54 | 0.024 | 0.030 | 4.29E-01 |
|  |  | MR RAPS | 54 | 0.035 | 0.018 | 5.25E-02 |
|  |  | Weighted median | 54 | 0.031 | 0.025 | 2.10E-01 |
|  |  | Weighted mode | 54 | 0.029 | 0.019 | 1.26E-01 |
| *CASP5* | CAD | Inverse variance weighted (fixed effects) | 28 | 0.000 | 0.024 | 1.00E+00 |
|  |  | MR Egger | 28 | 0.171 | 0.107 | 1.22E-01 |
|  |  | MR RAPS | 28 | 0.000 | 0.023 | 1.00E+00 |
|  |  | Weighted median | 28 | 0.038 | 0.033 | 2.47E-01 |
|  |  | Weighted mode | 28 | 0.039 | 0.031 | 2.17E-01 |
|  | Pneumonia | Inverse variance weighted (fixed effects) | 29 | -0.054 | 0.033 | 9.66E-02 |
|  |  | MR Egger | 29 | -0.004 | 0.080 | 9.59E-01 |
|  |  | MR RAPS | 29 | -0.055 | 0.033 | 9.57E-02 |
|  |  | Weighted median | 29 | -0.058 | 0.046 | 2.13E-01 |
|  |  | Weighted mode | 29 | -0.056 | 0.045 | 2.27E-01 |
| *DHX33* | CAD | Inverse variance weighted (fixed effects) | 4 | 0.179 | 0.068 | 9.06E-03 |
|  |  | MR Egger | 4 | 0.537 | 0.285 | 2.00E-01 |
|  |  | MR RAPS | 4 | 0.180 | 0.070 | 1.04E-02 |
|  |  | Weighted median | 4 | 0.226 | 0.084 | 7.00E-03 |
|  |  | Weighted mode | 4 | 0.230 | 0.095 | 9.36E-02 |
|  | Pneumonia | Inverse variance weighted (fixed effects) | 4 | 0.263 | 0.099 | 7.78E-03 |
|  |  | MR Egger | 4 | -0.112 | 0.542 | 8.56E-01 |
|  |  | MR RAPS | 4 | 0.266 | 0.101 | 8.64E-03 |
|  |  | Weighted median | 4 | 0.267 | 0.140 | 5.63E-02 |
|  |  | Weighted mode | 4 | 0.308 | 0.199 | 2.19E-01 |
| *IL4* | CAD | Inverse variance weighted (fixed effects) | 4 | 0.008 | 0.071 | 9.06E-01 |
|  |  | MR Egger | 4 | 0.430 | 0.294 | 2.81E-01 |
|  |  | MR RAPS | 4 | 0.009 | 0.071 | 9.03E-01 |
|  |  | Weighted median | 4 | 0.055 | 0.086 | 5.27E-01 |
|  |  | Weighted mode | 4 | 0.121 | 0.085 | 2.51E-01 |
|  | Pneumonia | Inverse variance weighted (fixed effects) | 4 | 0.101 | 0.093 | 2.78E-01 |
|  |  | MR Egger | 4 | 0.433 | 0.246 | 2.20E-01 |
|  |  | MR RAPS | 4 | 0.102 | 0.094 | 2.79E-01 |
|  |  | Weighted median | 4 | 0.175 | 0.099 | 7.65E-02 |
|  |  | Weighted mode | 4 | 0.176 | 0.108 | 2.00E-01 |
| *NEK7* | CAD | Inverse variance weighted (fixed effects) | 2 | 0.188 | 0.114 | 9.80E-02 |
|  | Pneumonia | Inverse variance weighted (fixed effects) | 2 | -0.078 | 0.164 | 6.35E-01 |
| *NLRC4* | CAD | Inverse variance weighted (fixed effects) | 22 | -0.046 | 0.027 | 9.10E-02 |
|  |  | MR Egger | 22 | -0.022 | 0.047 | 6.36E-01 |
|  |  | MR RAPS | 22 | -0.047 | 0.028 | 9.13E-02 |
|  |  | Weighted median | 22 | -0.041 | 0.033 | 2.21E-01 |
|  |  | Weighted mode | 22 | -0.036 | 0.032 | 2.80E-01 |
|  | Pneumonia | Inverse variance weighted (fixed effects) | 23 | -0.066 | 0.039 | 8.93E-02 |
|  |  | MR Egger | 23 | -0.115 | 0.083 | 1.81E-01 |
|  |  | MR RAPS | 23 | -0.066 | 0.039 | 8.88E-02 |
|  |  | Weighted median | 23 | -0.082 | 0.046 | 7.29E-02 |
|  |  | Weighted mode | 23 | -0.075 | 0.046 | 1.22E-01 |
| *NLRP3* | CAD | Inverse variance weighted (fixed effects) | 28 | -0.053 | 0.020 | 8.69E-03 |
|  |  | MR Egger | 28 | 0.020 | 0.042 | 6.38E-01 |
|  |  | MR RAPS | 28 | -0.054 | 0.020 | 8.62E-03 |
|  |  | Weighted median | 28 | -0.007 | 0.028 | 7.99E-01 |
|  |  | Weighted mode | 28 | -0.019 | 0.024 | 4.54E-01 |
|  | Pneumonia | Inverse variance weighted (fixed effects) | 28 | 0.083 | 0.029 | 3.57E-03 |
|  |  | MR Egger | 28 | 0.070 | 0.054 | 2.06E-01 |
|  |  | MR RAPS | 28 | 0.083 | 0.029 | 3.66E-03 |
|  |  | Weighted median | 28 | 0.087 | 0.037 | 1.73E-02 |
|  |  | Weighted mode | 28 | 0.083 | 0.033 | 1.88E-02 |
| *TXNIP* | CAD | single SNP | 1 | -0.072 | 0.123 | 5.61E-01 |
|  | Pneumonia | single SNP | 1 | 0.258 | 0.165 | 1.19E-01 |

###### eQTL data derived from whole blood for *IL6R, IL1B*, and *NLRP3* related genes were from the eQTLGen consortium. Mendelian randomization with either Wald ratio method (single instrument) or inverse-variance weighted, MR Egger, MR RAPS, weighted median, and weighted mode methods (more than one instrument) were used for this analysis. CAD: coronary artery disease, eQTL: expression quantitative trait loci.
